# Decreases in purchases of energy, sodium, sugar, and saturated fat three years after implementation of the Chilean Food Labelling and Marketing Law

**DOI:** 10.1101/2023.11.21.23298789

**Authors:** Lindsey Smith Taillie, Maxime Bercholz, Barry Popkin, Natalia Rebolledo, Marcela Reyes, M. Camila Corvalán

## Abstract

**Background:** In 2016, Chile implemented a multi-phase set of policies that mandated warning labels, restricted food marketing to children, and banned school sales of unhealthy foods and beverages. Chile’s law, particularly the warning label component, set the precedent for a rapid global proliferation of similar policies. While our initial evaluation showed policy-linked decreases in purchases of products carrying the warning label, a longer-term evaluation is needed, particularly as later phases of Chile’s law included stricter nutrient thresholds and introduced a daytime ban on advertising of labeled foods for all audiences. The objective is to evaluate changes in purchases of energy, sugar, sodium, and saturated fat purchased after Phase 2 implementation of the Chilean policies.

**Methods and Findings:** This before- and after-study used longitudinal data on monthly food and beverage purchases from 2,844 Chilean households (138,391 household-months) from July 1, 2013 until June 30, 2019. Nutrition facts panel data from food and beverage packages were linked at the product level and reviewed by nutritionists. Products were considered to carry the warning label if they contained added sugar, sodium, or saturated fat, and exceeded the final phase nutrient or calorie thresholds (thus would carry the warning label). Using correlated random-effects models and an interrupted time series design, we estimated the nutrient content of food and beverage purchases associated with Phase 1 and Phase 2 compared to a counterfactual scenario based on pre-policy trends. Compared to the counterfactual, we observed significant decreases in purchases of foods and beverages carrying the warning label during Phase 2, including a relative 36.8% reduction in sugar (−30.3 calories, 95% CI −34.5, −26.3), a 23.0% relative reduction in energy (−51.6 calories, 95% CI −60.7, −42.6), a 21.9% relative reduction in sodium (−85.8 mg, 95% CI −105.0, −66.7) and a 15.7% relative reduction in saturated fat (−6.4 calories, 95% CI −8.4, −4.3). Decreases were partially offset by increases in non-labeled purchases, but the net effect shows a significant decrease in total nutrients of concern purchased during Phase 2. Reductions in sugar and energy were driven by beverage purchases, whereas reductions in sodium and saturated fat were driven by foods. The pattern of declines in purchases was similar for households of lower vs. higher socioeconomic status. A key limitation of this study is that the data include only a portion of what Chilean households purchase that, while including important categories impacted by the law, do not cover an entire diet.

**Conclusions:** The Chilean policies on food labeling, marketing, and school food sales led to declines in nutrients of concern during a more complete phase of implementation, particularly from foods and drinks carrying the warning label.

## Introduction

In the last decade, many countries across the globe have taken action to halt increases in obesity prevalence and non-communicable disease risk by implementing policies designed to reduce consumption of unhealthy packaged foods and sugar-sweetened beverages^1^. In the years since Chile first began implementing its landmark policies on front-of-package labeling, restricted marketing, and ban of school sales of food and drinks high in calories, added sugar, sodium, and saturated fat (“high-in” foods), many countries around the globe have followed suit. Chile’s policy requiring mandatory warning labels on the front of “high-in” foods has seen particularly rapid momentum, with the majority of countries in South and North America now having implemented or in the process of implementing similar policies. Even in the United States, where policy progress has been slower, the 2022 National White House Conference on Hunger and Health recommended clearer front-of-package labels as an important strategy for improving public health nutrition, and the US Food and Drug Administration is now researching design options for a simple, interpretive front-of-package food label, including a version of warning labels.

The Chilean regulations were implemented in three phases (2016, 2018, and 2019). Initial evaluations of the Chilean policy after the first phase found that it was associated with changes in the food environment, including reductions of nutrients of concern in the food supply (e.g., energy, sugar, sodium, and saturated fat) ^2^ and reductions in children’s exposure to unhealthy food advertising and marketing^3–7^, improvements in mothers’ understanding of unhealthy foods and use of the warning labels^8^, reductions in purchases of foods and drinks carrying the warning label^9–11^, improvements in the nutritional quality of foods available in schools^12^, and improvement in children’s dietary intake^13^.

However, much less is known about policy-linked changes in dietary behavior over a longer time period. Evaluations of beverage taxes have shown sustained effects two years after policy implementation^14–16^, but to our knowledge, there have been no longer-term evaluations of a multi-component non-fiscal policy such as Chile’s. One concern is that the impact of certain parts of the law like the warning labels might wear off over time as consumers became accustomed to seeing the labels on packages, reducing the effect on food purchases. On the other hand, the second phase of Chile’s law, implemented in July 2018, included important changes to the regulations that could have increased the law’s impact on food purchases. Nutrient thresholds (particularly for foods) became stricter, meaning that more foods and beverages were affected by the regulations (i.e., more foods carried warning labels). In addition, the marketing restriction expanded to include the first-ever daytime ban on television advertising of ***all*** products carrying the warning labels to all audiences, not just those using child-directed appeals or appearing on children’s programs. While a recently published study found a 63% reduction in high-in food advertisements on television during Phase 2^17^, changes in food purchasing during this period are currently unclear.

Other questions relate to how the policy influenced specific product types. For example, it is unclear which food or beverage groups were most affected by the law, which is important for understanding nutrient changes and ultimately, dietary patterns. In addition, it is unclear the extent to which the policy shifted purchases from products with multiple warning labels to products with one warning label or no warning labels. This data is relevant to inform front-of-package labeling policies in other countries, who are grappling with the ideal number of warning labels to put on products in order to nudge consumers to healthier choices.

Finally, it is also important to understand whether the policies differentially influenced food purchases for households with low socioeconomic status (SES). A recent review of interpretive front-of-package labels found that while such labels are better understood than back-of-package nutrition information, relative to high-SES individuals, low-SES individuals are less likely to understand and use front-of-package labels and also less likely to shift purchasing intentions as a result of label exposure.^18^ In addition, food prices are a major driver of food choices in low-SES households^19^, potentially making them less likely to respond to non-fiscal policies like labeling and marketing policies. Understanding policy-linked purchasing changes among low-SES households is critical for ensuring future policies promote healthier diets for the entire population.

The objective of this study is to examine the pre-post association of the Chilean Law of Food Labeling and Advertising with food purchases during Phase 2 of implementation of the law. A secondary objective is to explore whether changes in purchases over time differed by household SES and by number of warning labels.

## Methods

This study was exempt from review by the University of North Carolina, Chapel Hill Institutional Review Board (IRB) as it used secondary, de-identified data. The study was approved by the University of Chile IRB.

### Study design and participants

We used longitudinal data on household food purchases from Kantar WorldPanel Chile from July 1, 2013 to June 25, 2019. Data were aggregated at the household-monthly level.

Households were excluded if they were missing demographic information for a given year. We also excluded all purchases in bulk (no quantity), purchases with a price or quantity of zero, baby food and formula, Kantar categories and subcategories that were introduced or discontinued during the study period as determined by changes in the number of purchases in these categories and subcategories over time (condensed milk, snacks, cereal bars, and minced meat), and household-month observations with total energy of zero. We also excluded household-month observations from new panelists before their first full month in the panel. We did so because Kantar Worldpanel began integrating new panelists around the last week of each month in February 2017, resulting in systematic measurement error in their first monthly totals. The final analytical sample included 2,844 unique households (median follow up, 64 months; 138,391 household-month observations) from cities (population > 20,000) across 13 of Chile’s 16 regions. The Kantar sample is weighted to be representative of these regions’ urban populations in terms of key demographics, and it is demographically similar to the overall Chilean population^10^.

Data on household purchases included the volume (ml) or weight (g), barcode, date, price, retail outlet, brand, and package size. Data were then linked at the product level to nutrition data, which was collected and updated annually from 2015 to 2019.

Data also included household characteristics, including size, composition (age and gender of each member), assets (number of rooms, bathrooms, and cars), geographical region, age of main shopper, head of household (primary earner) educational attainment, and socioeconomic status (SES). SES was defined by the market and public opinion research association Asociacion de Investigadores de Mercado y Opinion Publica de Chile and calculated based on household assets, head of household education, and access to goods and services^10^.

We further linked the data to the quarterly regional unemployment rate^20^ (downloaded from Chile’s statistics office’s website in September 2022) and public holiday data from the Python package ‘holidays’ (version 0.19, with minor corrections).

For main analyses, the pre-policy period was defined as July 1, 2013 to June 30, 2016, Phase 1 was defined as July 1, 2016 to June 30, 2018, and Phase 2 was defined as July 1, 2018 to June 30, 2019. Because Chile experienced national civil unrest in October 2019 followed by the Covid-19 pandemic in Feb-March of 2020, we were unable to include Phase 3 in this study (beginning July 1, 2019) due to the food supply disruptions.

### The Chilean regulation

Chile’s law requires that packaged foods and beverages containing added sugar, added sodium, or added saturated and exceeding thresholds for these nutrients or overall calories carry front-of-package warning labels. The labels consist of black octagon(s) with the text that it is high in sugar, sodium, saturated fat, and/or calories. The products are also subject to marketing restrictions (disallowed to use child-directed marketing techniques or air on platforms targeting children). The products are also banned from sales or promotion in schools and nurseries. The policy was implemented in phases with increasingly strict nutrient thresholds implemented in July 2016 (Phase 1); July 2018 (Phase 2); and July 2019 (Phase 3) (**Supplementary Table 1)**. The main difference between the Phase 1 and Phase 2 regulations was that for foods, the thresholds became much stricter for sodium (800 mg/100g to 500 mg/100g) and total sugars (22.5g to 15 g/100g), with smaller reductions for calories (350 calories to 300 calories per 100g) and saturated fat (6g/100g to 5g/100g). For beverages, changes in the thresholds were smaller and affected only calories (from 100 calories/100ml to 80 calories/100ml) and sugar (from 6 g/100ml to 5g/100ml). The other key difference between Phase 1 and Phase 2 is Phase 2 expanded the marketing restrictions to include a ban on advertising of products with the warning labels on daytime television from 6am to 10pm, regardless of audience (in other words, it was no longer just limited to children’s programming).

### Nutritional data and categorizations by food group and regulation status

As previously described, household purchases were linked at the product level to nutrition facts panel (NFP) data from food packages, Mintel Latin America, and other NFP resources using barcode, brand, and product description^21,22^. For the pre-policy period, purchases were linked to NFP data collected in the first quarters of 2015 and 2016; for Phase 1, purchases were linked to NFP data collected in the first quarters of 2017 and 2018; and for Phase 2, purchases were linked to NFP data collected in the first quarter of 2019.

Because the warning label serves as the visual signal as to whether the product is high in critical nutrients according to the Chilean regulation, we classified products as labeled or non-labeled based on their nutritional profile. Specifically, nutritionists reviewed each product for nutritional accuracy and, for consistency across all policy periods, applied the Phase 3 thresholds to categorize each product as subject to carry the warning label if it contained added sugar, sodium, or saturated fat and exceeded nutrient or calorie thresholds and thus was subject to the labeling, marketing, and school regulations.^23^ In other words, foods were classified as to whether they should receive the warning label under Chilean law, not whether they actually did (though compliance with the warning labels in Chile is >97%, so these are functionally the same). Foods were categorized as non-labeled if they did not exceed thresholds. Nutritionists also categorized foods into nutritionally and behaviorally relevant groups (**Supplementary Table 2**). Some groups were excluded from analysis because they were not affected by the regulation (e.g., plain teas, sweeteners, vegetable oils, formulas and supplements).

### Outcomes

All outcomes were estimated as per capita per day. As in our previous evaluation, the main outcome was total energy (calories), which was selected as the primary outcome due to its relevance for obesity prevention, one of the main goals of the law. We also examined total sugar (calories), saturated fat (calories), and sodium (mg). For all nutrients, we conducted analyses of overall purchases and for labeled and non-labeled purchases. Consistent with our first-phase evaluation^9^, we also examined food (all four outcomes) and beverage (energy and sugar) purchases separately and by subgroup (restricting our attention to subgroups accounting for at least 5% of aggregate food or beverage expenditure and with a relatively high proportion of high-in products).

### Statistical analysis

We used an interrupted time series design to estimate the changes in the nutritional content of food and beverage purchases associated with the first and second phases of the law. We assumed that each phase was associated with immediate proportionate changes and that these changes may have increased or decreased over time. These assumptions reflect both the nature of the intervention and the way we linked the purchase and nutrition data.

First, changes in amounts of energy, saturated fat, sodium, and sugar were likely proportional to their baseline levels rather than constant across all households. Second, because we linked the first 12 months of the first and second phases to nutrition data collected in the first quarter of 2017 and 2019, respectively, products that were reformulated in the first six to nine months of each phase are treated as if reformulation had been immediate. Third, we did not find evidence of widespread early or late adoption of the regulations before and after the law took effect. Finally, we allowed for changes in the impact of the law over time, albeit at a constant rate, for more flexibility.

We used correlated random-effects Poisson models together with the quasi-likelihood Poisson estimator with standard errors clustered at the household level to estimate the parameters of interest. The impact model took the form of a continuous variable for time (at monthly intervals), pre-post indicator variables for the first and second phases, and interactions with time. We controlled for seasonality by including indicator variables for each month of the year (reference: January), number of household members by age and sex (children aged 0 to 1 year, 2 to 5 years, and 6 to 13 years, females aged 14 to 18 years and 19 years or over, and males aged 14 to 18 years and 19 years or over), socio-economic status (ABC1, C2, C3, and DE; reference: ABC1), the head of household’s education level (less than high school, high school, and more than high school; reference: less than high school), the unemployment rate at the region and quarter level, the number of public holidays at the region and month level, and a pre-post indicator variable for the SSB tax implemented on October 1, 2014.

We then used the regression results to derive estimates of the average percentage and absolute changes associated with each phase of the law, calculated as differences in predicted values between the actual scenario and the counterfactual in which the law had not passed. We also estimated monthly means in the actual and counterfactual scenarios to depict these differences over time. We weighted these estimates using Kantar Worldpanel’s annual projection weights.

Separately, we created an indicator variable for low SES by combining the lowest two levels, and interacted it with each variable of the impact model to see if SES moderated the law’s impact. For these analyses, we focused on calories as our main outcome of interest. For these models, we dropped the head of household’s education level from the model because it is a component of SES, so keeping it would have meant restricting any moderation effect to the other components of SES.

To understand whether there were bigger changes for products that contained more labels (i.e., were high in more nutrients), we also examined the primary outcome by the number of warning labels. Finally, to understand if different food groups changed more or less after the law’s implementation, we conducted analyses using the same model as in the main analyses to examine nutrient outcomes for food and beverage subgroups.

### Sensitivity analyses

We conducted a series of sensitivities analyses. First, we reduced the length of the baseline period from 36 months (which balances the pre and post periods) to 24 and 18 months by dropping observations before January 2014 and July 2015, respectively. These analyses allow us to understand how the use of different baseline periods may affect results (either through changes to trends in purchasing behaviors, or changes to assumptions around reformulation, or both). Second, although the Poisson model is well-suited for non-negative skewed dependent variables, we compared our food and beverage group results to estimates obtained from a two-part model consisting of a correlated random-effects logit model in the first part (with the same explanatory variables) and the correlated random-effects Poisson model for the non-zero outcomes in the second part. All analyses were conducted in Stata 17 (College Station, TX, USA).

## Results

Descriptive characteristics of the sample can be found in **Table 1**. Household size increased over time; the household head educational attainment level also increased over time, while socioeconomic status remained consistent.

**Table 1.** Weighted socio-demographic characteristics of the Kantar WorldPanel sample by year.

|  | 2013 | 2014 | 2015 | 2016 | 2017 | 2018 | 2019 |
| --- | --- | --- | --- | --- | --- | --- | --- |
| Household size | 4.2<br>(1.6) | 4.4<br>(1.7) | 4.2<br>(1.6) | 4.2<br>(1.7) | 4.3<br>(1.7) | 4.4<br>(1.7) | 4.5<br>(1.7) |
| Main shopper's age | 47.9<br>(14.5) | 48.3<br>(14.7) | 48.2<br>(15.0) | 48.6<br>(14.9) | 49.3<br>(15.2) | 49.4<br>(15.5) | 49.6<br>(15.5) |
| Number of cars | 0.5<br>(0.7) | 0.6<br>(0.7) | 0.7<br>(0.8) | 0.7<br>(0.7) | 0.7<br>(0.7) | 0.7<br>(0.7) | 0.7<br>(0.8) |
| Number bathrooms | 1.3<br>(0.6) | 1.3<br>(0.6) | 1.3<br>(0.6) | 1.3<br>(0.6) | 1.3<br>(0.6) | 1.3<br>(0.6) | 1.3<br>(0.7) |
| Number bedrooms | 2.9<br>(1.0) | 3.0<br>(2.4) | 3.1<br>(1.5) | 3.0<br>(1.0) | 3.0<br>(1.0) | 3.0<br>(1.0) | 3.1<br>(1.0) |
| Household head's education |  |  |  |  |  |  |  |
| Lower than high school | 42% | 40% | 37% | 32% | 31% | 28% | 27% |
| High school | 37% | 38% | 40% | 42% | 42% | 45% | 44% |
| Higher than high school | 21% | 22% | 23% | 25% | 27% | 28% | 29% |
| Socioeconomic status (SES) |  |  |  |  |  |  |  |
| ABC1 (highest) | 8% | 8% | 8% | 8% | 8% | 8% | 7% |
| C2 | 17% | 17% | 17% | 16% | 16% | 17% | 16% |
| C3 | 27% | 27% | 27% | 27% | 27% | 27% | 28% |
| DE (lowest) | 49% | 49% | 49% | 49% | 49% | 48% | 49% |
| Region |  |  |  |  |  |  |  |
| North | 13% | 13% | 12% | 13% | 13% | 13% | 13% |
| Center | 64% | 63% | 63% | 63% | 63% | 62% | 62% |
| South | 24% | 24% | 24% | 24% | 24% | 25% | 25% |
| Number of household-month obs. | 11,636 | 23,455 | 23,292 | 23,385 | 22,762 | 22,684 | 11,177 |

Notes: The sample starts in July 2013 and ends in June 2019. The definition of SES is based on Chile's Association for Market Research and Public Opinion (*Asociacion de Investigadores de Mercado y Opinion Publica*), which is a measure of socioeconomic status based on a list of goods and services consumed by the household, the education level of the head of household, and income.

Prior to the law, approximately 44% of foods were eligible to carry 2 or more warning labels, 25% were eligible to carry one warning label, and 31% would have had zero warning labels (**Supplementary Table 3)**. For beverages, only 3% were eligible to carry two or more warning labels, 39% were eligible to carry one warning label, and 58% would have had zero warning labels.

### Unadjusted results

Unadjusted results on household purchases can be found in **Supplemental Tables 4 and 5**. The pattern of results showed a decline in nutrients of concern purchased across the study time period as well as declines in the share of nutrients from purchases of products with the warning label (“labeled products”), ranging from −5.6 percentage points for saturated fat to −16.5 percentage points for sugar.

### Adjusted results

Comparing weighted estimates of purchases as observed to the counterfactual scenario, we observe significant decreases in purchases of labeled foods and beverages during Phase 2 (**Figure 1**), including a relative 36.8% reduction in sugar (−30.4 calories, 95% CI −34.5, −26.3), a 23.0% relative reduction in energy (−51.6 calories, 95% CI −60.7, −42.6), a 21.9% relative reduction in sodium (−85.8 mg, 95% CI −105.0, −66.7) and lastly a 15.7% relative reduction in saturated fat (−6.4 calories, 95% CI −8.4, −4.3). These decreases were partially compensated by increases in non-labeled purchases (**Table 2**), but the net effect shows a significant decrease in total nutrients purchased during Phase 2, resulting in relative reductions of 20.2% in sugar, 13.8% in sodium, −9.6% in saturated fat, and 8.3% in energy from total food and beverage purchases. The pattern of results was similar between Phase 1 and Phase 2.

**Figure 1.**
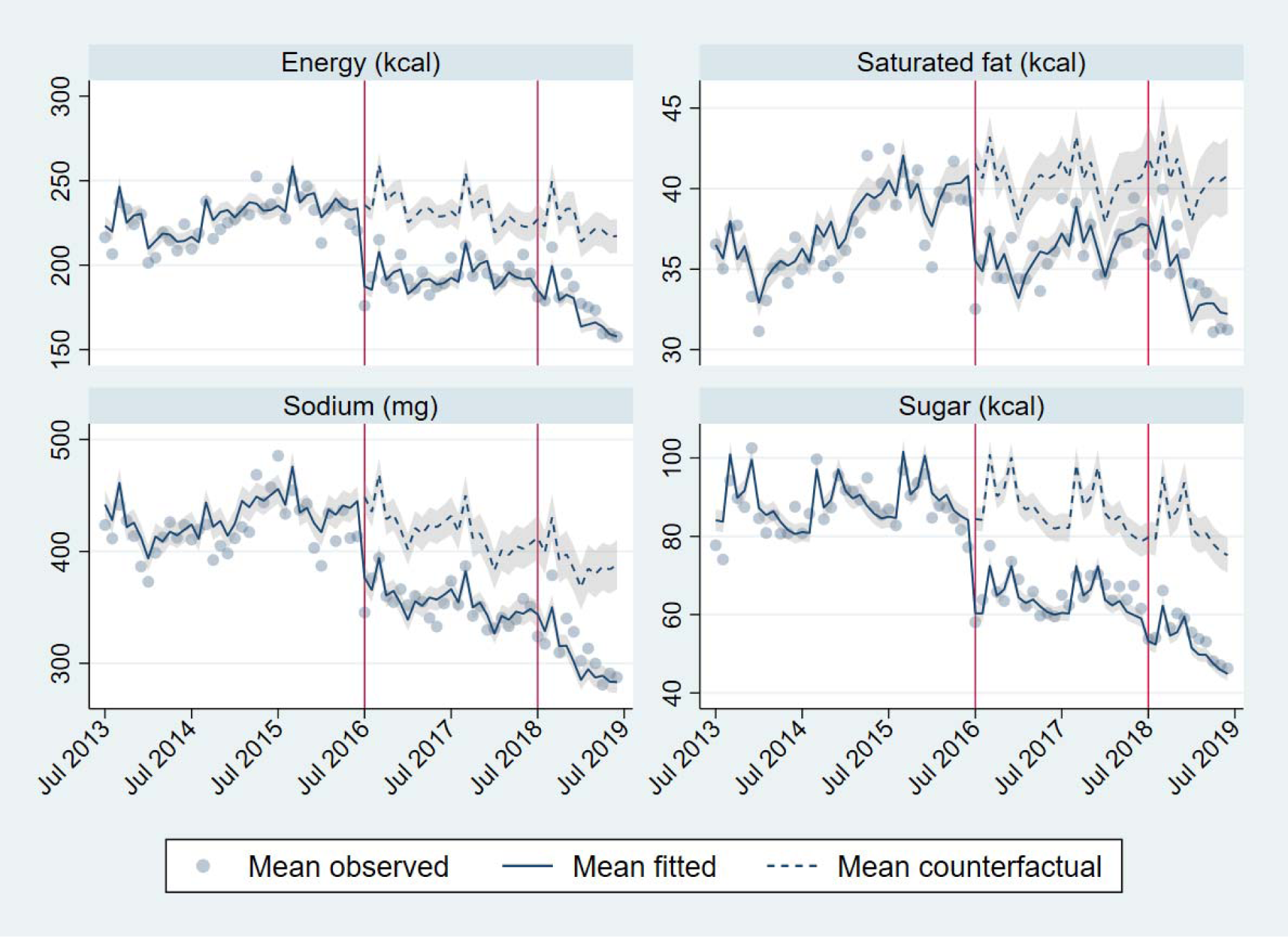
Mean adjusted weighted purchases of nutrients from labeled food and beverage purchases during the pre-policy period, Phase 1, and Phase 2 of implementation of the Chilean Law of Food Labeling and Advertising

**Table 2.**
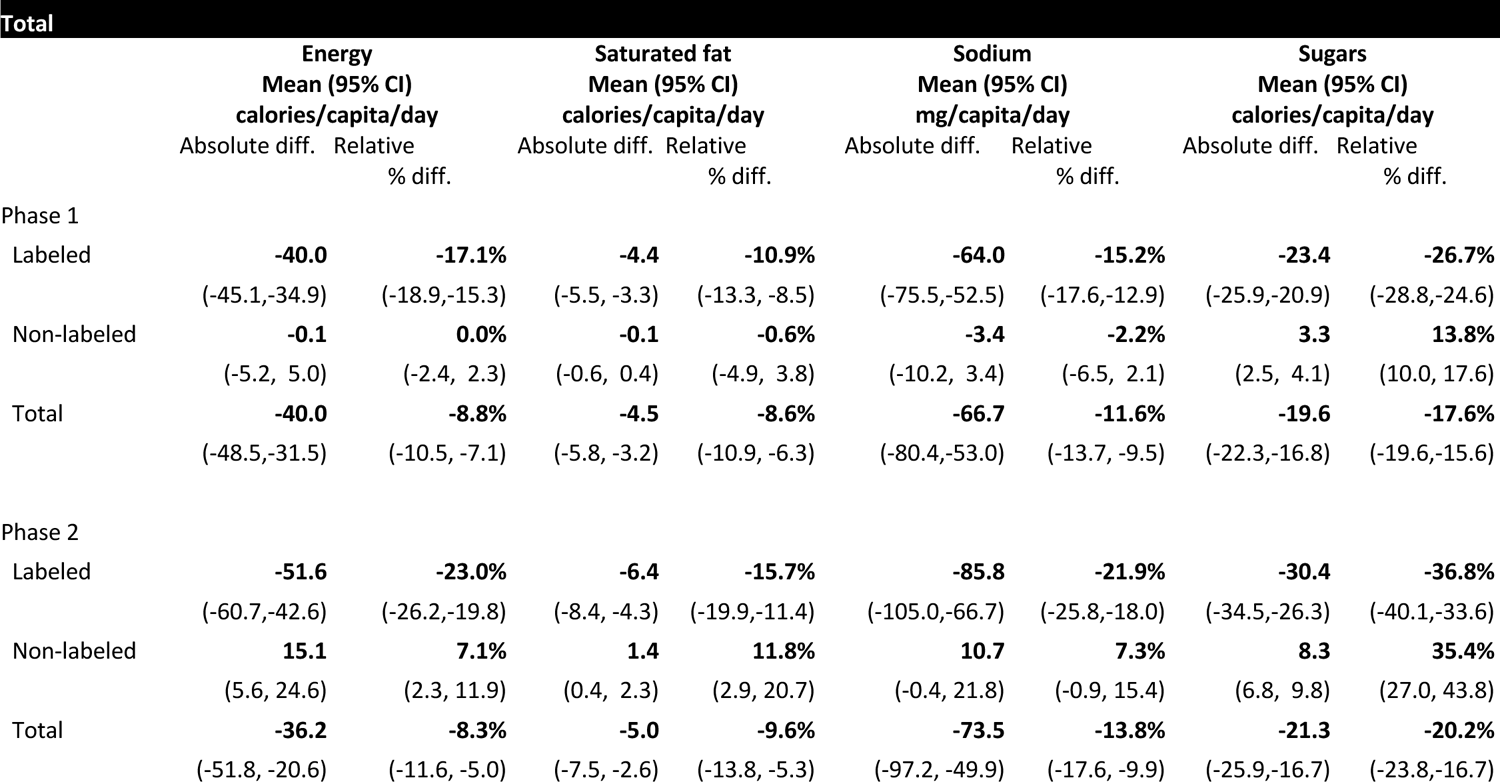
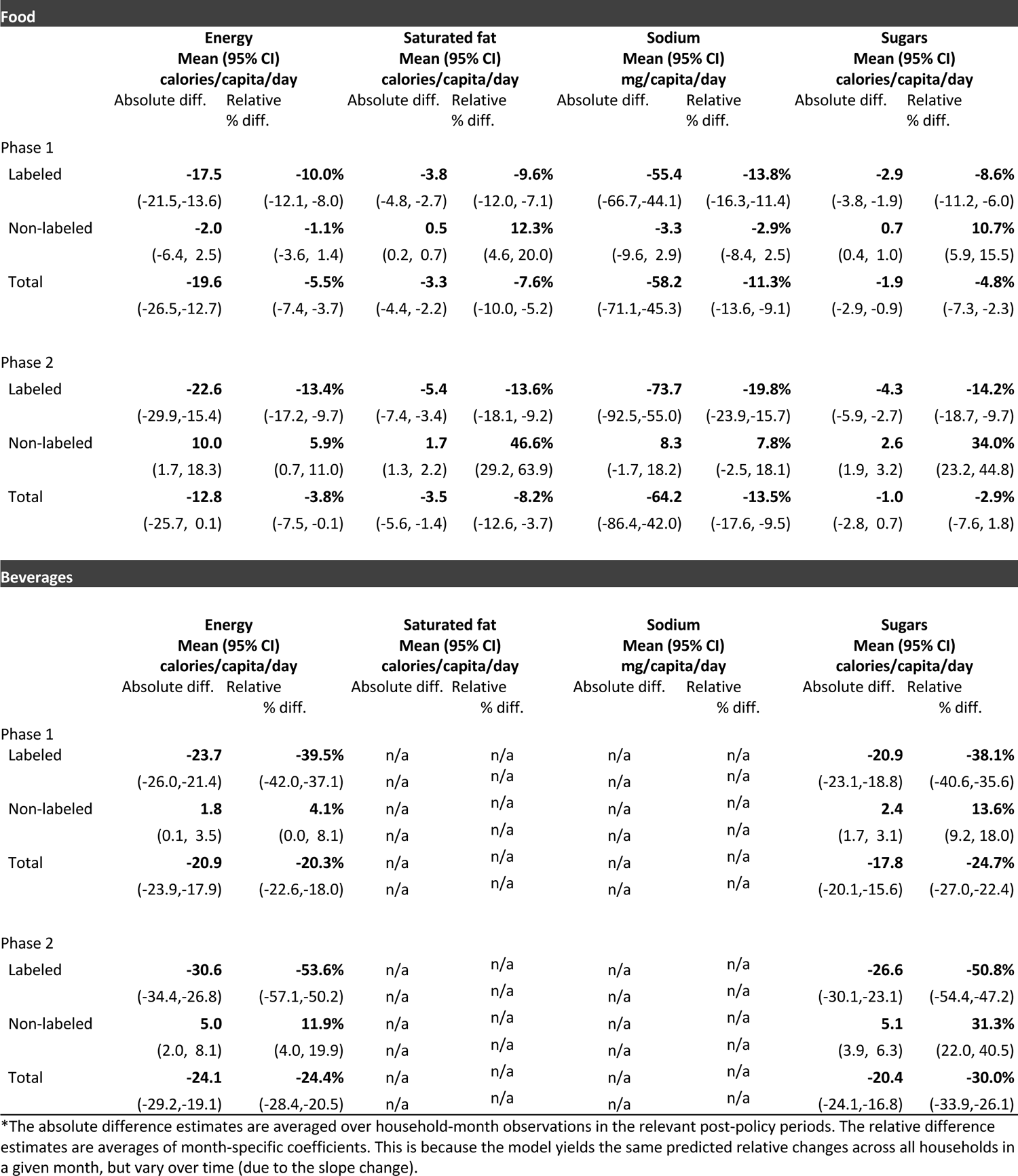
Adjusted weighted mean differences in nutrient content of total purchases, food purchases, and beverage purchases between the estimated adjusted Phase 1 and Phase 2 post-policy periods and the counterfactual scenario.

| Total |  |  |  |  |  |  |  |  |
| --- | --- | --- | --- | --- | --- | --- | --- | --- |
|  | Energy<br>Mean (95% CI)<br>calories/capita/day |  | Saturated fat<br>Mean (95% CI)<br>calories/capita/day |  | Sodium<br>Mean (95% CI)<br>mg/capita/day |  | Sugars<br>Mean (95% CI)<br>calories/capita/day |  |
|  | Absolute diff. | Relative<br>% diff. | Absolute diff. | Relative<br>% diff. | Absolute diff. | Relative<br>% diff. | Absolute diff. | Relative<br>% diff. |
| Phase 1 |  |  |  |  |  |  |  |  |
| Labeled | **-40.0**<br>(-45.1,-34.9) | **-17.1%**<br>(-18.9,-15.3) | **-4.4**<br>(-5.5, -3.3) | **-10.9%**<br>(-13.3, -8.5) | **-64.0**<br>(-75.5,-52.5) | **-15.2%**<br>(-17.6,-12.9) | **-23.4**<br>(-25.9,-20.9) | **-26.7%**<br>(-28.8,-24.6) |
| Non-labeled | **-0.1**<br>(-5.2, 5.0) | **0.0%**<br>(-2.4, 2.3) | **-0.1**<br>(-0.6, 0.4) | **-0.6%**<br>(-4.9, 3.8) | **-3.4**<br>(-10.2, 3.4) | **-2.2%**<br>(-6.5, 2.1) | **3.3**<br>(2.5, 4.1) | **13.8%**<br>(10.0, 17.6) |
| Total | **-40.0**<br>(-48.5,-31.5) | **-8.8%**<br>(-10.5, -7.1) | **-4.5**<br>(-5.8, -3.2) | **-8.6%**<br>(-10.9, -6.3) | **-66.7**<br>(-80.4,-53.0) | **-11.6%**<br>(-13.7, -9.5) | **-19.6**<br>(-22.3,-16.8) | **-17.6%**<br>(-19.6,-15.6) |
| Phase 2 |  |  |  |  |  |  |  |  |
| Labeled | **-51.6**<br>(-60.7,-42.6) | **-23.0%**<br>(-26.2,-19.8) | **-6.4**<br>(-8.4, -4.3) | **-15.7%**<br>(-19.9,-11.4) | **-85.8**<br>(-105.0,-66.7) | **-21.9%**<br>(-25.8,-18.0) | **-30.4**<br>(-34.5,-26.3) | **-36.8%**<br>(-40.1,-33.6) |
| Non-labeled | **15.1**<br>(5.6, 24.6) | **7.1%**<br>(2.3, 11.9) | **1.4**<br>(0.4, 2.3) | **11.8%**<br>(2.9, 20.7) | **10.7**<br>(-0.4, 21.8) | **7.3%**<br>(-0.9, 15.4) | **8.3**<br>(6.8, 9.8) | **35.4%**<br>(27.0, 43.8) |
| Total | **-36.2**<br>(-51.8, -20.6) | **-8.3%**<br>(-11.6, -5.0) | **-5.0**<br>(-7.5, -2.6) | **-9.6%**<br>(-13.8, -5.3) | **-73.5**<br>(-97.2, -49.9) | **-13.8%**<br>(-17.6, -9.9) | **-21.3**<br>(-25.9,-16.7) | **-20.2%**<br>(-23.8,-16.7) |

| Food |  |  |  |  |  |  |  |  |
| --- | --- | --- | --- | --- | --- | --- | --- | --- |
|  | Energy<br>Mean (95% CI)<br>calories/capita/day |  | Saturated fat<br>Mean (95% CI)<br>calories/capita/day |  | Sodium<br>Mean (95% CI)<br>mg/capita/day |  | Sugars<br>Mean (95% CI)<br>calories/capita/day |  |
|  | Absolute diff. | Relative<br>% diff. | Absolute diff. | Relative<br>% diff. | Absolute diff. | Relative<br>% diff. | Absolute diff. | Relative<br>% diff. |
| Phase 1 |  |  |  |  |  |  |  |  |
| Labeled | **-17.5**<br>(-21.5,-13.6) | **-10.0%**<br>(-12.1, -8.0) | **-3.8**<br>(-4.8, -2.7) | **-9.6%**<br>(-12.0, -7.1) | **-55.4**<br>(-66.7,-44.1) | **-13.8%**<br>(-16.3,-11.4) | **-2.9**<br>(-3.8, -1.9) | **-8.6%**<br>(-11.2, -6.0) |
| Non-labeled | **-2.0**<br>(-6.4, 2.5) | **-1.1%**<br>(-3.6, 1.4) | **0.5**<br>(0.2, 0.7) | **12.3%**<br>(4.6, 20.0) | **-3.3**<br>(-9.6, 2.9) | **-2.9%**<br>(-8.4, 2.5) | **0.7**<br>(0.4, 1.0) | **10.7%**<br>(5.9, 15.5) |
| Total | **-19.6**<br>(-26.5,-12.7) | **-5.5%**<br>(-7.4, -3.7) | **-3.3**<br>(-4.4, -2.2) | **-7.6%**<br>(-10.0, -5.2) | **-58.2**<br>(-71.1,-45.3) | **-11.3%**<br>(-13.6, -9.1) | **-1.9**<br>(-2.9, -0.9) | **-4.8%**<br>(-7.3, -2.3) |
| Phase 2 |  |  |  |  |  |  |  |  |
| Labeled | **-22.6**<br>(-29.9,-15.4) | **-13.4%**<br>(-17.2, -9.7) | **-5.4**<br>(-7.4, -3.4) | **-13.6%**<br>(-18.1, -9.2) | **-73.7**<br>(-92.5,-55.0) | **-19.8%**<br>(-23.9,-15.7) | **-4.3**<br>(-5.9, -2.7) | **-14.2%**<br>(-18.7, -9.7) |
| Non-labeled | **10.0**<br>(1.7, 18.3) | **5.9%**<br>(0.7, 11.0) | **1.7**<br>(1.3, 2.2) | **46.6%**<br>(29.2, 63.9) | **8.3**<br>(-1.7, 18.2) | **7.8%**<br>(-2.5, 18.1) | **2.6**<br>(1.9, 3.2) | **34.0%**<br>(23.2, 44.8) |
| Total | **-12.8**<br>(-25.7, 0.1) | **-3.8%**<br>(-7.5, -0.1) | **-3.5**<br>(-5.6, -1.4) | **-8.2%**<br>(-12.6, -3.7) | **-64.2**<br>(-86.4,-42.0) | **-13.5%**<br>(-17.6, -9.5) | **-1.0**<br>(-2.8, 0.7) | **-2.9%**<br>(-7.6, 1.8) |
| Beverages |  |  |  |  |  |  |  |  |
|  | Energy<br>Mean (95% CI)<br>calories/capita/day |  | Saturated fat<br>Mean (95% CI)<br>calories/capita/day |  | Sodium<br>Mean (95% CI)<br>mg/capita/day |  | Sugars<br>Mean (95% CI)<br>calories/capita/day |  |
|  | Absolute diff. | Relative<br>% diff. | Absolute diff. | Relative<br>% diff. | Absolute diff. | Relative<br>% diff. | Absolute diff. | Relative<br>% diff. |
| Phase 1 |  |  |  |  |  |  |  |  |
| Labeled | **-23.7**<br>(-26.0,-21.4) | **-39.5%**<br>(-42.0,-37.1) | n/a | n/a | n/a | n/a | **-20.9**<br>(-23.1,-18.8) | **-38.1%**<br>(-40.6,-35.6) |
| Non-labeled | **1.8**<br>(0.1, 3.5) | **4.1%**<br>(0.0, 8.1) | n/a | n/a | n/a | n/a | **2.4**<br>(1.7, 3.1) | **13.6%**<br>(9.2, 18.0) |
| Total | **-20.9**<br>(-23.9,-17.9) | **-20.3%**<br>(-22.6,-18.0) | n/a | n/a | n/a | n/a | **-17.8**<br>(-20.1,-15.6) | **-24.7%**<br>(-27.0,-22.4) |
| Phase 2 |  |  |  |  |  |  |  |  |
| Labeled | **-30.6**<br>(-34.4,-26.8) | **-53.6%**<br>(-57.1,-50.2) | n/a | n/a | n/a | n/a | **-26.6**<br>(-30.1,-23.1) | **-50.8%**<br>(-54.4,-47.2) |
| Non-labeled | **5.0**<br>(2.0, 8.1) | **11.9%**<br>(4.0, 19.9) | n/a | n/a | n/a | n/a | **5.1**<br>(3.9, 6.3) | **31.3%**<br>(22.0, 40.5) |
| Total | **-24.1**<br>(-29.2,-19.1) | **-24.4%**<br>(-28.4,-20.5) | n/a | n/a | n/a | n/a | **-20.4**<br>(-24.1,-16.8) | **-30.0%**<br>(-33.9,-26.1) |

\*The absolute difference estimates are averaged over household-month observations in the relevant post-policy periods. The relative difference estimates are averages of month-specific coefficients. This is because the model yields the same predicted relative changes across all households in a given month, but vary over time (due to the slope change).

We observe that nutrient reductions are more pronounced for beverages than for foods for sugar and calories. For example, in Phase 2, there was a 53.6% relative reduction in energy from labeled beverages (−30.6 calories, 95% CI −34.4, −26.8), but only a 13.4% relative reduction in energy from labeled foods (−22.6 calories, 95% CI −29.9,-15.4); and only for beverages did these reductions in labeled energy purchases translate to significant reductions in total energy purchases, with a similar pattern for sugar purchases. However, among food purchases, there were significant reductions in both sodium and saturated fat purchases, both among labeled and total food purchases.

The pattern of results by number of warning labels also differed for foods and beverages. For foods, we observed reductions in calories only for foods that had at least two warning labels, whereas in Phase 2, calories purchased increased for foods that had only one warning (**Table 3**). In contrast, we observed large reductions among beverages that carried a single warning and only small reductions among beverages with two or more warnings (likely because very few beverages contain more than one warning).

**Table 3.** Adjusted weighted mean differences in energy content of total purchases, food purchases, and beverage purchases between the estimated adjusted Phase 1 and Phase 2 post-policy periods and the counterfactual scenario, by number of warning labels.

| **Total** |  |  |  |  |
| --- | --- | --- | --- | --- |
|  | **Phase 1**<br>**Mean (95% CI)**<br>**calories/capita/day** |  | **Phase 2**<br>**Mean (95% CI)**<br>**calories/capita/day** |  |
|  | Absolute diff. | Relative % diff. | Absolute diff. | Relative % diff. |
| No warning labels | **-0.1** | **0.0%** | **15.1** | **7.1%** |
|  | (-5.2, 5.0) | (-2.4, 2.3) | (5.6, 24.6) | (2.3, 11.9) |
| 1 warning label | **-16.8** | **-17.6%** | **-22.7** | **-25.7%** |
|  | (-19.6, -14.0) | (-20.1, -15.1) | (-27.2, -18.1) | (-29.6, -21.7) |
| ≥2 warning labels | **-23.4** | **-16.9%** | **-29.5** | **-21.6%** |
|  | (-26.9, -19.9) | (19.0, -14.8%) | (-36.0, -23.1) | (-25.4, -17.9) |
| All | **-40.0** | **-8.8%** | **-36.2** | **-8.3%** |
|  | (-48.5, -31.5) | (-10.5, -7.1) | (-51.8, -20.6) | (-11.6, -5.0) |
| **Food** |  |  |  |  |
|  | **Phase 1**<br>**Mean (95% CI)**<br>**calories/capita/day** |  | **Phase 2**<br>**Mean (95% CI)**<br>**calories/capita/day** |  |
|  | Absolute diff. | Relative % diff. | Absolute diff. | Relative % diff. |
| No warning labels | **-2.0** | **-1.1%** | **10.0** | **5.9%** |
|  | (-6.4, 2.5) | (-3.6, 1.4) | (1.7, 18.3) | (0.7, 11.0) |
| 1 warning label | **3.2** | **8.7%** | **4.2** | **11.8%** |
|  | (1.9, 4.6) | (3.8, 12.7) | (1.9, 6.4) | (3.7, 18.8) |
| ≥2 warning labels | **-20.8** | **-15.3%** | **-27.1** | **-20.3%** |
|  | (-24.2, -17.4) | (-17.4, -13.2) | (-33.4, -20.8) | (-24.0, -16.5) |
| All | **-19.6** | **-5.5%** | **-12.8** | **-3.8%** |
|  | (-26.5, -12.7) | (-7.4, -3.7) | (-25.7, 0.1) | (-7.5, -0.1) |
| **Beverages** |  |  |  |  |
|  | **Phase 1**<br>**Mean (95% CI)**<br>**calories/capita/day** |  | **Phase 2**<br>**Mean (95% CI)**<br>**calories/capita/day** |  |
|  | Absolute diff. | Relative<br>% diff. | Absolute diff. | Relative<br>% diff. |
| No warning labels | **1.8** | **4.1%** | **5.0** | **11.9%** |
|  | (0.1, 3.5) | (0.0, 8.1) | (2.0, 8.1) | (4.0, 19.9) |
| 1 warning label | **-20.9** | **-36.7%** | **-28.0** | **-51.6%** |
|  | (-23.1, -18.6) | (-39.3, -34.1) | (-31.7, -24.3) | (-55.2, -48.0) |
| ≥2 warning labels | **-2.6** | **-89.4%** | **-2.4** | **-91.8%** |
|  | (-3.1, -2.1) | (-91.3, -87.4) | (-3.1, -1.7) | (-96.3, -87.3) |
| All | **-20.9** | **-20.3%** | **-24.1** | **-24.5%** |
|  | (23.9, -17.9) | (-22.6, -18.0) | (-29.2, -19.1) | (-28.4, -20.5) |

### Differences by SES Groups

Compared to their respective counterfactuals, there were few differences in absolute or relative declines in calories from total, food, or beverage purchases by socio-economic status (low vs. high) in either Phase 1 or Phase 2 of the regulation (**Table 4**). The exception to this is that for beverages, the relative decline in calories purchased from labeled beverages was greater for high SES vs. low SES households in both phases of the law, possibly due to high-SES households’ having lower purchases of labeled beverages during the pre-policy period (**Supplementary Table 5)**.

**Table 4.**
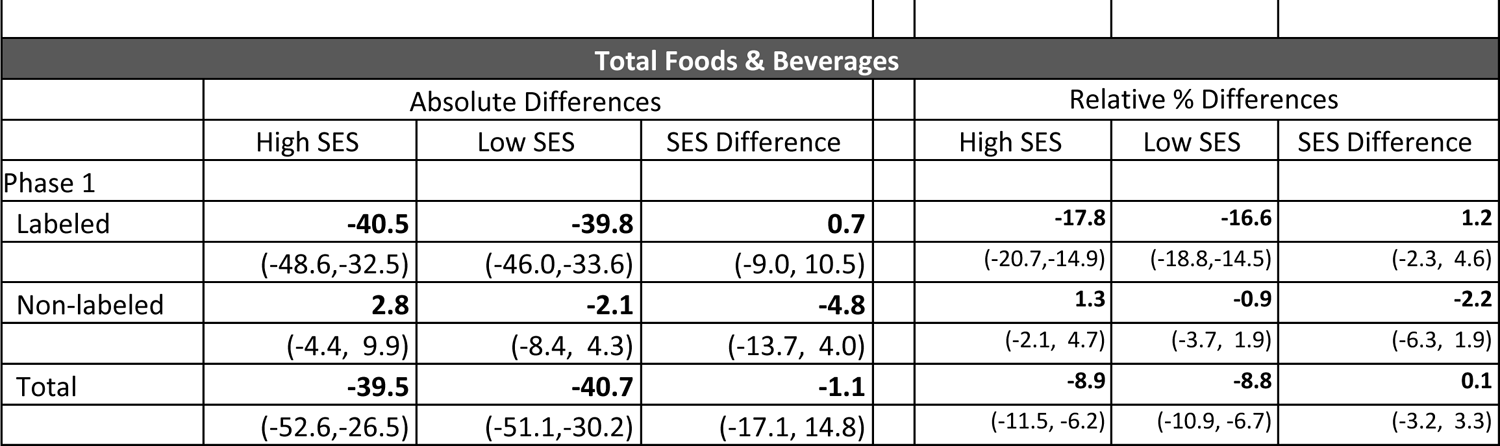
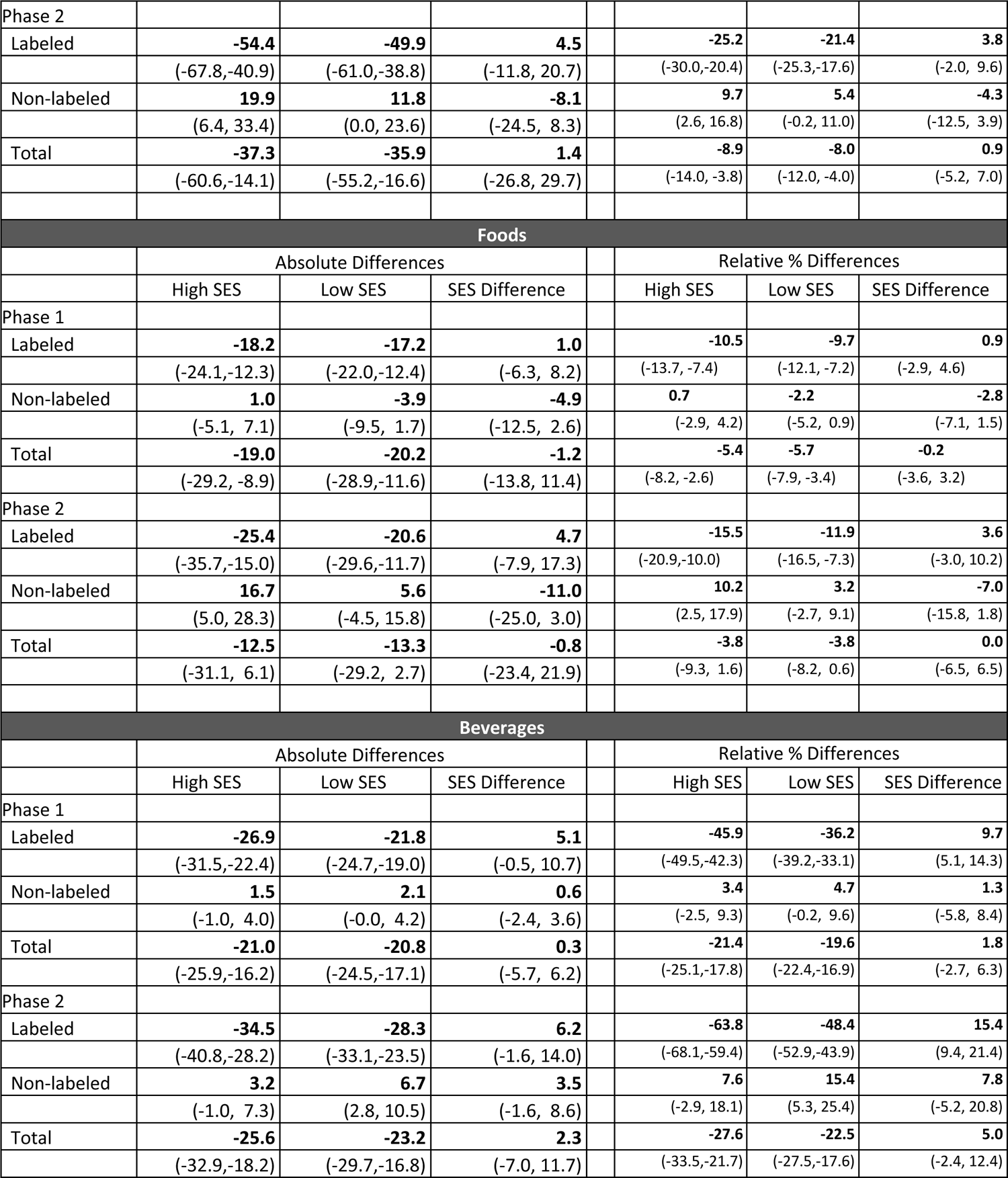
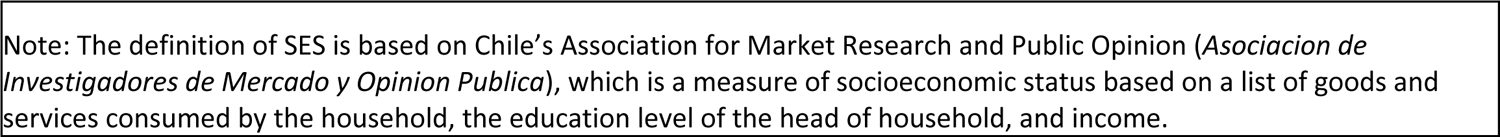
Adjusted mean differences in energy content of total food and beverage purchases between the Phase 1 and Phase 2 post-policy periods and the counterfactual scenario by socioeconomic status (SES), Mean Difference (95% CI)

### Food and Beverage Group Adjusted Results

Food and beverage subgroup results can be found in **Table 5 and Table 6**, respectively.

**Table 5.** Adjusted mean differences in nutrient content of food group purchases between the Phase 1 and Phase 2 post-policy periods and the counterfactual scenario, Mean Difference (95% CI)

|  | Calories |  |  |  | Sugar |  |  |  |
| --- | --- | --- | --- | --- | --- | --- | --- | --- |
|  | Phase 1 |  | Phase 2 |  | Phase 1 |  | Phase 2 |  |
|  | Absolute difference | % difference | Absolute difference | % difference | Absolute difference | % difference | Absolute difference | % difference |
| Breakfast cereals | -1.8 | -12.1 | -1.9 | -12.5 | -0.6 | -21.4 | -0.8 | -27.8 |
|  | (-2.7, -0.9) | (-17.7, -6.5) | (-3.6, -0.2) | (-22.5, -2.5) | (-0.8, -0.4) | (-27.2, -15.7) | (-1.1, -0.4) | (-37.5, -18.1) |
| Grain-based desserts | -2.6 | -10.8 | -2.1 | -8.4 | -0.7 | -10.6 | -0.6 | -8.8 |
|  | (-3.8, -1.4) | (-15.1, -6.4) | (-4.4, 0.1) | (-16.4, -0.3) | (-1.0, -0.4) | (-14.9, -6.2) | (-1.2, 0.0) | (-16.8, -0.7) |
| Sweets and non-grain-based desserts | -1.5 | -6.3 | -0.9 | -4.2 | -0.8 | -5.8 | -0.4 | -3.1 |
|  | (-2.4, -0.5) | (-10.4, -2.2) | (-2.5, 0.7) | (-11.4, 3.1) | (-1.4, -0.2) | (-9.9, -1.7) | (-1.3, 0.6) | (-10.4, 4.1) |
| Meat, poultry and meat substitutes | -2.1 | -6.4 | -2.4 | -7.7 | -0.2 | -33.0 | -0.1 | -18.3 |
|  | (-3.2, -0.9) | (-9.8, -2.9) | (-4.5, -0.3) | (-13.9, -1.5) | (-0.3, -0.2) | (-36.8, -29.3) | (-0.2, -0.1) | (-26.2, -10.4) |
| Dairy products and substitutes | -0.8 | -3.6 | -1.5 | -7.1 | -0.1 | -1.5 | -0.2 | -3.0 |
|  | (-1.7, 0.2) | (-7.9, 0.7) | (-3.3, 0.2) | (-14.9, 0.6) | (-0.5, 0.2) | (-6.3, 3.2) | (-0.9, 0.4) | (-11.5, 5.6) |
| Condiments and sauces | -0.7 | -2.8 | 1.3 | 5.9 | -0.3 | -7.6 | -0.4 | -8.5 |
|  | (-1.6, 0.3) | (-6.8, 1.3) | (-0.4, 2.9) | (-2.0, 13.8) | (-0.5, -0.2) | (-11.2, -4.0) | (-0.7, -0.1) | (-14.8, -2.2) |
| Oils and fats | -3.4 | -9.4 | -4.1 | -11.3 | 0.0 | 40.0 | 0.0 | 47.6 |
|  | (-4.8, -2.0) | (-12.9, -5.8) | (-6.8, -1.5) | (-17.7, -4.8) | (0.0, 0.0) | (31.3, 48.6) | (0.0, 0.0) | (32.5, 62.6) |
|  | Saturated Fat |  |  |  | Sodium |  |  |  |
|  | Phase 1 |  | Phase 2 |  | Phase 1 |  | Phase 2 |  |
|  | Absolute difference | % difference | Absolute difference | % difference | Absolute difference | % difference | Absolute difference | % difference |
| Breakfast cereals | -0.1 | -15.3 | -0.1 | -9.8 | -1.5 | -18.8 | -2.6 | -32.5 |
|  | (-0.1, -0.0) | (-21.8, -8.8) | (-0.1, 0.0) | (-22.2, 2.6) | (-2.1, -0.9) | (-24.8, -12.8) | (-3.7, -1.5) | (-41.8, -23.2) |
| Grain-based desserts | -0.6 | -11.9 | -0.6 | -11.8 | -1.7 | -12.6 | -1.6 | -11.5 |
|  | (-0.8, -0.3) | (-16.5, -7.4) | (-1.0, -0.1) | (-20.1, -3.6) | (-2.3, -1.0) | (-17.1, -8.2) | (-2.8, -0.3) | (-19.6, -3.4) |
| Sweets and non-grain-based desserts | -0.3 | -10.1 | -0.6 | -21.1 | 0.0 | -0.1 | 0.2 | 1.9 |
|  | (-0.5, -0.1) | (-15.3, -4.8) | (-0.9, -0.3) | (-29.3, -12.8) | (-0.5, 0.4) | (-4.9, 4.7) | (-0.6, 0.9) | (-6.6, 10.5) |
| Meat, poultry and meat substitutes | -0.4 | -4.9 | -0.5 | -6.6 | -22.8 | -18.3 | -20.9 | -17.6 |
|  | (-0.6, -0.1) | (-8.4, -1.3) | (-1.0, 0.0) | (-13.0, -0.3) | (-27.5, -18.2) | (-21.3, -15.3) | (-29.1, -12.7) | (-23.3, -11.8) |
| Dairy products and substitutes | -0.3 | -4.7 | -0.4 | -7.9 | -1.9 | -12.1 | -4.2 | -26.9 |
|  | (-0.5, 0.0) | (-9.8, 0.4) | (-1.0, 0.1) | (-17.2, 1.4) | (-2.7, -1.2) | (-16.3, -8.0) | (-5.5, -2.8) | (-33.4, -20.5) |
| Condiments and sauces | -0.2 | -7.1 | 0.1 | 4.2 | -13.1 | -14.7 | -15.0 | -17.6 |
|  | (-0.3, -0.0) | (-12.1, -2.0) | (-0.1, 0.3) | (-5.9, 14.2) | (-16.2, -10.0) | (-17.6, -11.8) | (-20.3, -9.7) | (-22.7, -12.4) |
| Oils and fats | -1.5 | -8.4 | -1.5 | -8.5 | -3.9 | -10.1 | -8.4 | -23.0 |
|  | (-2.2, -0.7) | (-12.2, -4.5) | (-2.9, -0.1) | (-15.7, -1.3) | (-5.4, -2.3) | (-13.8, -6.4) | (-11.2, -5.7) | (-28.8, -17.1) |

**Table 6.** Adjusted mean differences in nutrient content of beverage group purchases between the Phase 1 and Phase 2 post-policy periods and the counterfactual scenario, Mean Difference (95% CI)

|  | Calories |  |  |  | Sugar |  |  |  |
| --- | --- | --- | --- | --- | --- | --- | --- | --- |
|  | Phase 1 |  | Phase 2 |  | Phase 1 |  | Phase 2 |  |
|  | Absolute difference | % difference | Absolute difference | % difference | Absolute difference | % difference | Absolute difference | % difference |
| Sodas | -8.3 | -20.9 | -9.5 | -25.9 | -8.0 | -20.9 | -9.2 | -25.6 |
|  | (-10.0, -6.6) | (-24.4, -17.5) | (-12.3, -6.6) | (-32.0, -19.9) | (-9.7, -6.4) | (-24.4, -17.4) | (-12.0, -6.3) | (-31.7, -19.5) |
| Fruit and vegetable juice | -5.9 | -54.3 | -8.2 | -66.4 | -6.0 | -58.5 | -8.3 | -70.2 |
|  | (-6.7, -5.1) | (-57.3, -51.3) | (-9.7, -6.7) | (-70.5, -62.3) | (-6.8, -5.2) | (-61.3, -55.7) | (-9.7, -6.8) | (-73.9, -66.6) |
| Dairy-based drinks* | -7.4 | -16.0 | -8.8 | -19.2 | -4.4 | -21.8 | -4.9 | -25.1 |
|  | (-9.2, -5.5) | (-19.3, -12.6) | (-12.1, -5.5) | (-25.1, -13.3) | (-5.2, -3.6) | (-24.8, -18.7) | (-6.3, -3.5) | (-30.5, -19.7) |

\*Includes plant-based substitutes

For foods, across both phases, calories purchased from breakfast cereals; meat, poultry, and meat substitutes; and oils and fats declined, whereas calories from grain-based desserts, sweets and non-grain-based desserts only declined during Phase 1, and there were no changes in calories from dairy products/substitutes or condiments and sauces in either phase. Across all food groups and across both phases, absolute declines in sugar were trivial (less than one calorie/capita/day). Similarly, declines in saturated fat purchased were also trivial. Sodium purchases declined in both phases for every food group except grain-based desserts. The largest reductions were observed among meat, poultry, and meat substitutes (−20.9 mg/capita/day in Phase 2, 95% CI −29.1, −12.7, or a 17.6% reduction), condiments and sauces (−15.0 mg/capita/day in Phase 2, 95% CI −20.3, −9.7, or a 17.6% reduction), and oils and fats (−8.4 mg/capita/day in Phase 2, 95% CI −11.2, −5.7, or a 23.0% reduction).

For beverages, across both phases and across all groups, both calories and sugar purchases declined. Declines in calories and sugar in all beverage groups analyzed were larger than declines in calories and sugar in any food group. For both calories and sugar, the biggest declines were observed in sodas. There was an 8.3 calorie/capita/day reduction in soda calories purchased in Phase 1 (95% CI −10.0, −6.6), or a 20.9% reduction, and a 9.5 calorie/capita/day reduction in soda calories purchased in Phase 2 (95% CI −12.3, −6.6), or a 25.9% reduction; these reductions in calories were almost entirely driven by reductions in sugar. Declines in calories were similar for fruit and vegetable juices and for dairy-based beverages and substitutes (8.2 and 8.8 calories/capita/day in Phase 2, respectively), whereas declines in sugar were larger for juices (−8.3 calories/capita/day in Phase 2, 95% CI −9.7, −6.8) than for dairy-based beverages (−4.9, 95% CI −6.3, −3.5).

### Sensitivity analyses

Results from models run using different pre-policy windows on total, labeled, and not labeled calories from foods and beverages can be found in **Supplementary Table 6**. Results for total calorie changes during Phase 1 were generally consistent across from 24 to 42 months; however, reductions in total calories were smaller for the model that used only an 18 month time window. There was more variability in Phase 2 total calorie changes, though again the results consistently reflected a decline in total calories purchased when pre-policy windows from 24 to 42 months were used. However, there was no change in total calories when only an 18 month pre-policy window was used. This lack of effect in total calories during Phase 2 using an 18 month pre-policy window is because the relative increase in non-labeled calories got larger, while the decrease in labeled calories got smaller, resulting in no net impact on total calories. Results using two-part models for food and beverage subgroups were consistent with our main analysis **Supplementary Table 7**.

## Discussion

The second phase of Chile’s law of food labeling and advertising continued the warning label, child-directed marketing, and school foods policies, tightened nutrient thresholds for foods subject to these policies, and imposed a daytime ban on all unhealthy food advertisements on television, regardless of audience. After implementation of this second phase, we observed declines in purchases of nutrients of concern from food and drinks carrying the warning label (and thus subject to all regulations), including a 36.8% decline in sugar, 23.0% decline in energy, 21.9% decline in sodium, and 15.7% decline in saturated fat purchased. The declines from labeled products purchased were partially offset by increased purchases from non-labeled products. Still, the overall changes resulted in net declines in nutrients of concerned purchase ranging between 8.3% to 20.2%.

Declines in purchases of labeled foods and drinks were larger in Phase 2 than in Phase 1, as would be expected based on the stricter nutrient thresholds and daytime marketing ban that begin in Phase 2. At the same time, however, there were also larger increases in purchases of not labeled foods and beverages in Phase 2, resulting in a partial attenuation on the total effect on nutrient purchases (e.g., for total calories). While longer-term data from food policy evaluations are scarce, research from tobacco suggests that some attenuation of results over time is to be expected from marketing and labeling policies. One particular concern is message fatigue, in which long-term exposure to health-related messages like Chile’s warning labels leads to boredom or lack of motivation^24,25^. Indeed, Chilean mothers have reported paying less attention to the warning labels over time, and that as the prevalence of warning labels increased (as a function of stricter nutrient thresholds), they began to feel oversaturated with labels and also that the labels were not providing new information^26^. We should note here that Chile, unlike some of the more recent countries with warning labels like Mexico and Colombia, Chile did not have active civil society groups that continued to promote the warning label. After an initial government promotion campaign at the beginning of the warning labels, no future campaigns in the media existed, which may have influenced continued attention to and use of the warning labels.

Other aspects of the Chilean policy, like the daytime ban of television advertising of labeled foods introduced in Phase 2, may simply need longer to work, as research in this area suggests that the most likely immediate impacts of such a policy are changes to food attitudes and preferences,^27^ which may take years to translate into behavioral changes. In other words, the removal of food advertisements does not immediately negate prior exposure to years or decades of food marketing. In addition, industry adaptations may further attenuate these effects. For example, while there was a 64% drop in labeled food advertisements during Phase 2^16^, 29% of food ads promoted a labeled product, and 34% of children’s exposure to food ads was exposure to labeled ads^17^. Additionally, although there was a 14% decrease in total food ads, there was continued advertising of non-labeled products and brands,^17^ which could increase purchases.

Moreover, there has been a trend towards increased expenditures on digital food advertising in Chile^28^ and while digital advertisements are regulated by the Chilean law on child-directed marketing, they are not subject to the daytime ban, which is only for television, and overall monitoring, enforcement, and compliance in the digital space is unclear. These shifts in advertising could work to maintain consumerś preferences given that previous research suggests that advertising a particular product has the potential to increase preferences for the advertised product, its product category, and brand^29–31^. Furthermore, marketing impact consumerś brand knowledge, attitudes, and attachment which wońt change immediately after a reduction of marketing exposure^32–34^.

We observed differential results for foods vs. beverages across policy periods. For example, while declines in total energy were similar for foods and beverages in Phase 1, in Phase 2, the estimated overall decline for beverages was nearly double that of foods. This difference is attributable both to a larger drop in labeled purchases as well as a smaller compensation in not high in purchases for beverages compared to foods. With regards to total sugar, there was essentially no change in sugar from food purchases in either phase, whereas the decrease in sugar calories from beverage purchases was sizeable in both Phases due to the large drop in labeled beverages. These results are surprising given that foods were much more affected by stricter nutrient thresholds introduced in Phase 2 (e.g., the sugar threshold for foods went from 22.5 g/100 g in Phase 1 to 15 g/100g in Phase 2, whereas for beverages, the sugar threshold dropped only from 6g/100ml in Phase 1 to 5g/100ml in Phase 2).

There are several explanations for the differential results on sugar and energy for foods vs. beverages. One likely possibility is that beverages are easier to reformulate than are foods, since replacing sugar with non-nutritive sweeteners (NNS) may be more likely to cause issues with texture or taste in food products than in drinks^35–38^. Although data from Phase 2 are not yet available, data on the Chilean food supply from Phase 1 showed that NNS use was most prevalent among beverages before the law as well as the highest absolute increase after the law of any food category.^39^ Another evaluation found that, compared to the counterfactual, purchases of NNS in beverages, but not in foods, also increased after Phase 1 of the law^37^. Notably, multiple other countries who have implemented warning label laws after Chile’s have also included a warning for the presence of NNS. It will be important to understand how policy-linked changes in purchases of sugar and NNS compare in these countries as opposed to Chile, which did not include an NNS warning label, particularly for beverage purchases.

A second difference between foods and beverages relates to the number of warning labels on products. Because foods are more nutritionally diverse, they are more likely to be high in multiple nutrients of concern and thus more likely to contain multiple warning labels. We observed that among foods, there were decreases in calories purchased among foods that contained 2 or more warning labels, but increases in calories purchased for foods that contained one or no warning labels. This suggests that there is a tendency for consumers to shift purchases away from products with multiple warning labels towards those with fewer warnings, either through changes in consumer behavior (selecting a product with 1 vs 2 or more labels) or reformulation (companies reducing the nutrient content so as to reduce the total number of warning labels on the package). In contrast, among beverages it is much more common for products to carry only one warning label, usually on sugar content. Consequently, we observed calorie reductions for purchases of products carrying a single warning label. Taken together, the results suggest an overall shift from more labels to fewer labels: for foods, the shift is from products with multiple warning labels to those with fewer warning labels, whereas for beverages the shift is from products with a single warning label to those with no warning labels.

A third difference between foods and beverages is that that the policies were associated with important declines in sodium purchases from foods, whereas for beverages we did not analyze sodium because so few products were high in them. Purchases of sodium from labeled foods declined by approximately 74 mg/sodium/capita/day, or a nearly 20% relative reduction, and these decreases led to a net reduction of 64 mg/capita/day of total sodium purchases. Sodium reductions occurred across every food group except grain-based desserts, and were particularly sizeable for meat, poultry, and meat substitutes, condiments and sauces, and oils and fats. These results are encouraging, considering that Chile is one of only 5% of the 194 WHO member countries to achieve the highest score for sodium reduction policies, which includes having at least two mandatory policies, all WHO sodium related “best buy” practices, and sodium declarations on packaged foods^40^. Our results suggest that such policies lead to reductions in sodium purchases, though more research is needed to understand how these changes translate to achieving dietary targets for sodium in the Chilean population as well as subsequent sodium-related health benefits such as reduction of high blood pressure and cardiovascular disease. Overall, the results across sugar, sodium, and saturated fat highlight the need to include both foods and beverages in policies in order to reduce nutrients of concern.

The result that low-SES households reduced purchases of nutrients of concern after policy implementation were consistent with other data from our evaluation. Our focus group data found that low-SES parents paid attention to, understood, and used the warning labels^8,41^. In addition, a recent study found that during Phase 1 of the policy implementation, there were no price changes for labeled or not-high in foods^42^. Such price changes, for example if non-labeled-food prices increased as the result of companies passing on the cost of reformulation, could have dampened the effect among low-SES households who tend to be more price sensitive. Instead, the lack of price changes likely increased low-SES parents’ ability to attend to and incorporate information from the warning labels into their decision making.

However, a key limitation of our current study is that it did not include data after June 2019, a period which was marked by Chilean social unrest, food supply chain volatility, global food price inflation, and the Covid-19 pandemic and lockdowns, events that could have blunted the effect of the policies in low-SES households. Our qualitative data from 2021 suggested that even though the labels helped parents understand which foods were healthy vs. unhealthy, low-SES parents were struggling to choose the healthy options due to cost. Thus, more research is needed to understand the ways in which marketing, labeling, and school foods policies intersect with food prices, particularly during times of economic instability and global food price increases. Moreover, it is important to recognize that price remains a top driver of food choice, particularly for income-constrained households, and marketing and labeling policies to disincentivize unhealthy food choices may have limited effect if not implemented alongside policies that increase affordability or availability of healthier food choices.

## Limitations

This study had important limitations. One limitation is that the data included present only a portion of what Chilean households purchase, including important categories that may have been affected by the law (e.g., salty snacks) as well as loose, raw produce and other minimally processed foods that are not typically included in datasets on packaged food purchases (e.g., bread from a bakery). Beyond purchasing data, data on total dietary intake is necessary to fully evaluate the policies, particularly given that one of the policies focused on school food sales, data that is not captured in Kantar WorldPanel. Another key limitation is that while the construction of a counterfactual allowed us to compare observed purchases in the post-policy period to what would have happened in the absence of the policy, decisions about how to create the counterfactual are somewhat arbitrary and can influence results. We conducted sensitivity analyses using different policy windows to construct the counterfactual and showed that the pattern of results remains consistent, though effect sizes can vary. We also provide a more in-depth comparison of current findings with our team’s previous findings in **Supplementary Table 8**^43^.

One key limitation to the study is that we could not, as originally planned, evaluate changes in food purchases during Phase 3 of the Chilean law. Phase 3 was implemented in June of 2019. In the following year, the food supply and food purchasing behaviors were severely impacted by national protests in October-December 2019 and lockdowns due to the onset of the Covid-19 pandemic, making it impossible to analyze purchasing trends during this period. However, the results in this study are likely similar to what we would have observed during Phase 3. Although the nutrient thresholds became slightly stricter in Phase 2, there was only a 2% increase in the prevalence of labeled products, and there were no other substantive policy changes during this time. Of course, we cannot rule out other changes that could have occurred during Phase 3 (e.g., more consumer fatigue), but generally, the results presented here for Phase 2 are likely to represent the full implementation of the Chilean law.

## Data Availability

All data produced in the present study are available upon reasonable request to the authors.

## Acknowledgments

While Bloomberg Philanthropies provided much of the funding for this work, earlier grants from the Chilean government (CONICYT) and the Canadian IDRC were also important in starting this effort. We thank Fernanda Mediano for her insights on interpreting the marketing aspects of the Chilean law. We thank Bridget Hollingsworth for overall project management and Donna Miles for data management. We also thank research assistants, including Cindy Granados Evans, Fernanda Olivato, Jessica Ostrowski, Leonela Muñoz Conolly, Julia Wandell, Michelle Perry, Catalina Cornejo Nuñez, and Gina Bautista Herrera, as well as programmers, including Neepa Boode and Stephanie Stewart.

## Supplemental Figures

**Supplementary Table 1.** Nutrient thresholds and implementation dates of the Chilean Labeling and Advertising Law.

| **Solid food** | **26 June 2016** | **26 June 2018** | **26 June 2019** |
| --- | --- | --- | --- |
| Energy<br>(kcal/100g) | 350 | 300 | 275 |
| Sodium<br>(mg/100g) | 800 | 500 | 400 |
| Total sugars<br>(g/100g) | 22.5 | 15 | 10 |
| Saturated fats<br>(g/100g) | 6 | 5 | 4 |
| **Liquids** | **26 June 2016** | **26 June 2018** | **26 June 2019** |
| Energy<br>(kcal/100g) | 100 | 80 | 70 |
| Sodium<br>(mg/100g) | 100 | 100 | 100 |
| Total sugars<br>(g/100g) | 6 | 5 | 5 |
| Saturated fats<br>(g/100g) | 3 | 3 | 3 |

**Supplemental Table 2.** Food and beverage categories included and their aggregate expenditure shares.

| **Category** | **Food**<br>**% expenditure** | **Beverages**<br>**% expenditure** | **Total**<br>**% expenditure** |
| --- | --- | --- | --- |
| Cereal-based foods | 22.1 |  | 13.6 |
| Meat, poultry and meat substitutes* | 15.7 |  | 9.6 |
| Dairy products and dairy substitutes* | 13.1 |  | 8.0 |
| Sweets and non-grain-based desserts* | 12.2 |  | 7.5 |
| Condiments and sauces* | 10.2 |  | 6.3 |
| Oils and fats* | 8.1 |  | 5.0 |
| Grain-based desserts* | 7.2 |  | 4.4 |
| Breakfast cereals* | 5.0 |  | 3.1 |
| Soups | 3.3 |  | 2.0 |
| Snacks | 2.0 |  | 1.2 |
| Traditional mixed dishes | 0.5 |  | 0.3 |
| Salt and seasoning | 0.3 |  | 0.2 |
| Fruits, vegetables and mushrooms | 0.2 |  | 0.1 |
| Fish and seafood | 0.0 |  | 0.0 |
| Total food | 100.0 |  | 61.4 |
| Dairy-based beverages and dairy substitutes* |  | 37.5 | 14.5 |
| Sodas* |  | 33.4 | 13.0 |
| Industrialized, fruit and vegetable juice* |  | 9.9 | 3.8 |
| Water |  | 9.7 | 3.8 |
| Coffee and tea |  | 9.3 | 3.6 |
| 100% fruit/vegetable juice |  | 0.1 | 0.0 |
| Sports and energy drinks |  | 0.0 | 0.0 |
| Total beverages |  | 100.0 | 38.7 |
| Total |  |  | 100.0 |

Note: asterisks indicate categories that had $\geq 5\%$ share of expenditures in food or in beverages and were included in the food and beverage sub-group analyses.

**Supplemental Table 3.** Raw percentages of products with zero, one, and two or more FOPLs (Phase 3 limits) by period (July 2013-June 2019)

|  | Pre-policy | Phase 1 | Phase 2 |
| --- | --- | --- | --- |
| Food |  |  |  |
| None | 30.8 | 34.8 | 38.6 |
| One | 24.9 | 26.5 | 24.2 |
| Two or more | 44.2 | 38.7 | 37.2 |
| Beverages |  |  |  |
| None | 58.4 | 76.2 | 82.7 |
| One | 39.0 | 22.5 | 16.8 |
| Two or more | 2.5 | 1.2 | 0.5 |
| Total |  |  |  |
| None | 39.3 | 47.2 | 51.9 |
| One | 29.3 | 25.3 | 22.0 |
| Two or more | 31.4 | 27.5 | 26.1 |

As a result of the NFP linking protocol, the number of FOPLs a product has may vary within a given period (multiple years of NFP data used in the pre-policy and phase 1 periods).

**Supplemental Table 4.**
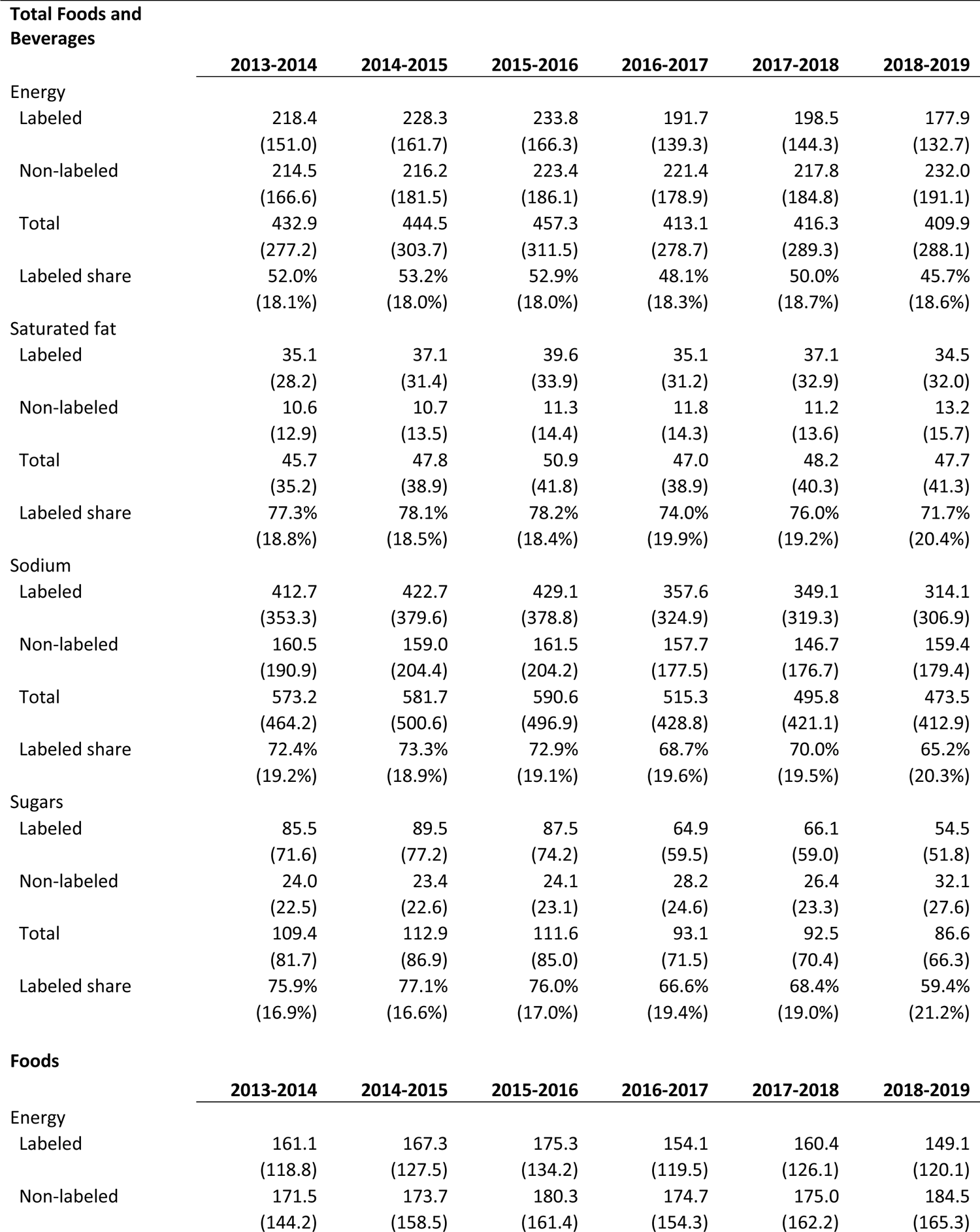
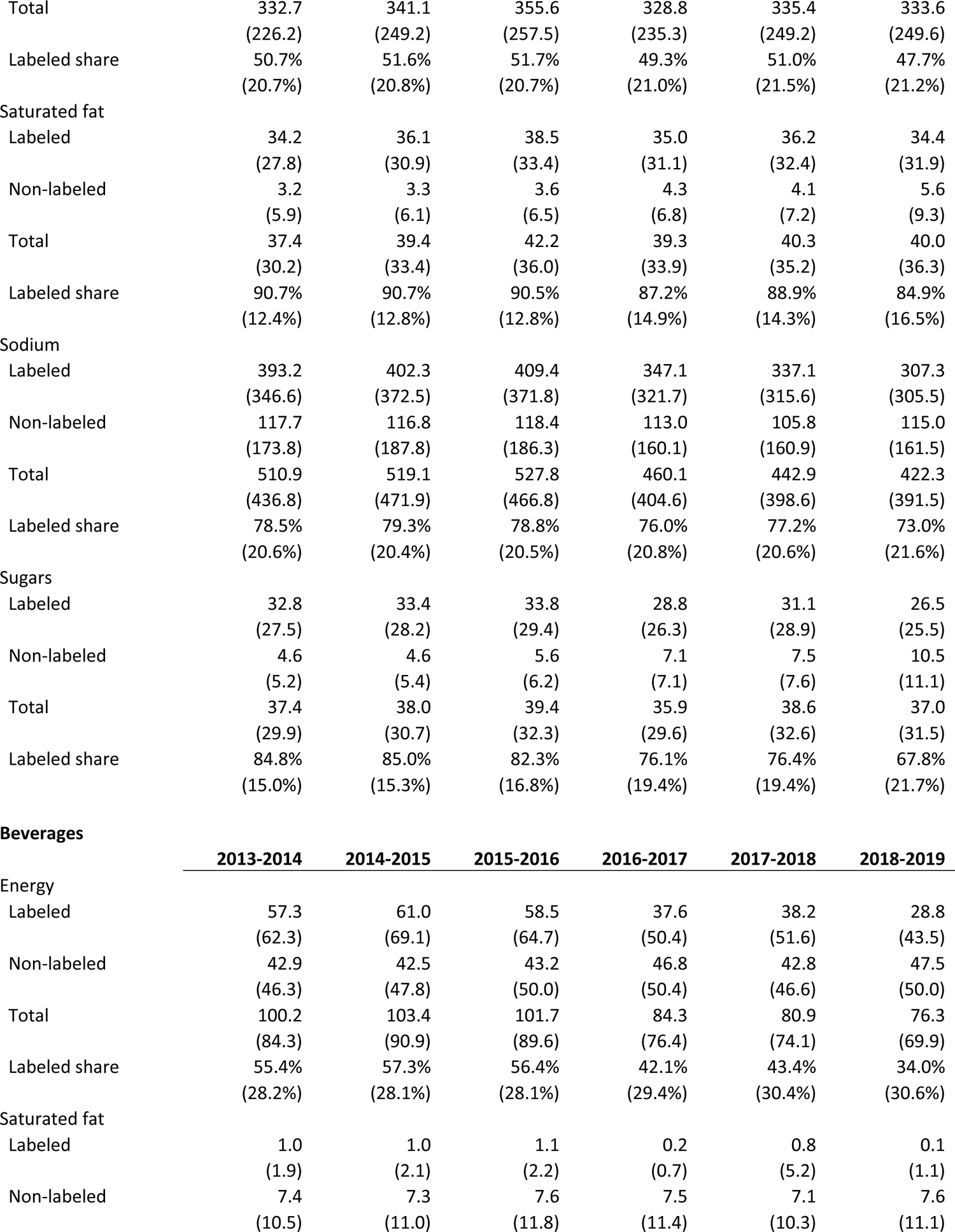
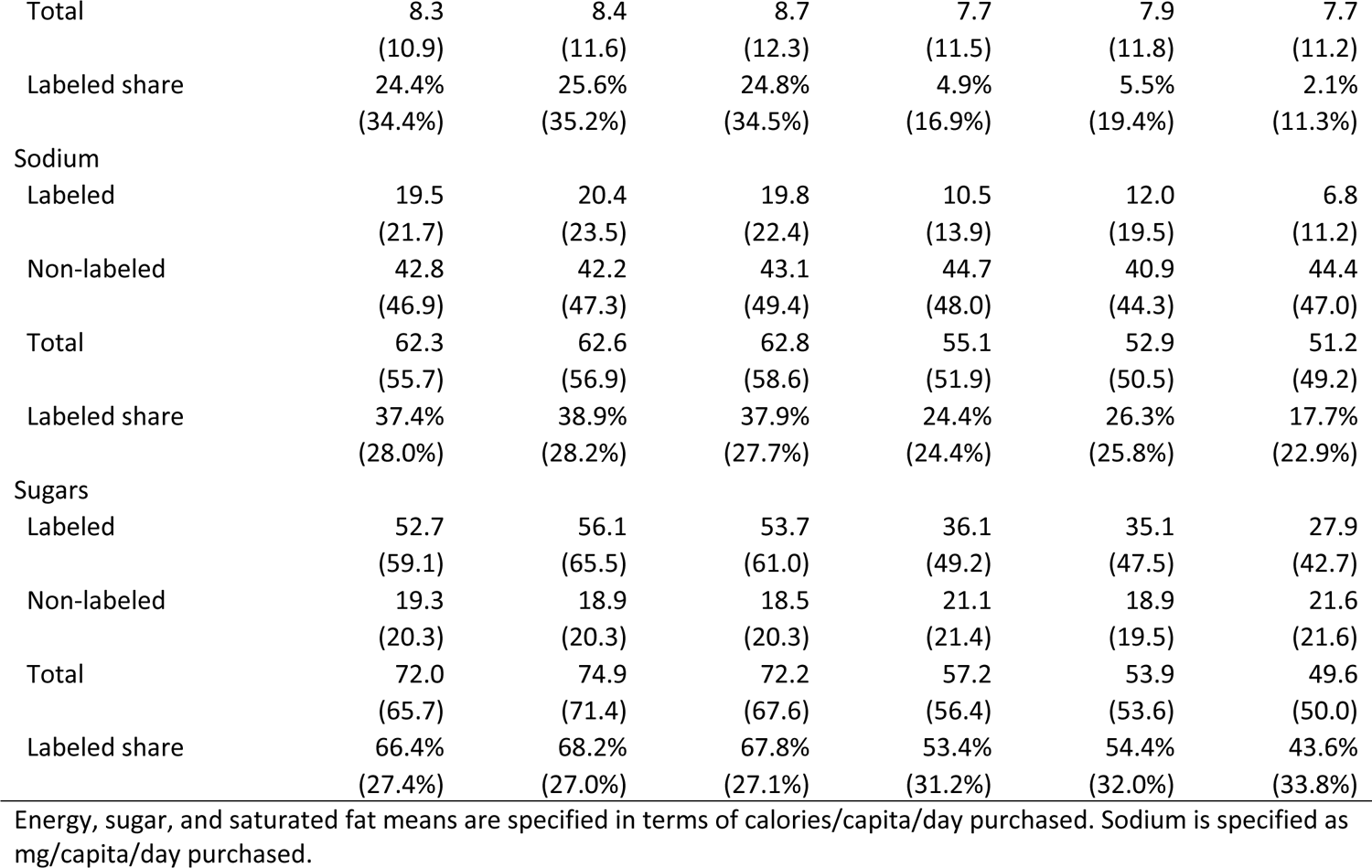
Unadjusted weighted means (SD) in food and beverage purchases by July-June period.

| **Total Foods and Beverages** |  | **2013-2014** | **2014-2015** | **2015-2016** | **2016-2017** | **2017-2018** | **2018-2019** |
| --- | --- | --- | --- | --- | --- | --- | --- |
| Energy |  |  |  |  |  |  |  |
| Labeled |  | 218.4<br>(151.0) | 228.3<br>(161.7) | 233.8<br>(166.3) | 191.7<br>(139.3) | 198.5<br>(144.3) | 177.9<br>(132.7) |
| Non-labeled |  | 214.5<br>(166.6) | 216.2<br>(181.5) | 223.4<br>(186.1) | 221.4<br>(178.9) | 217.8<br>(184.8) | 232.0<br>(191.1) |
| Total |  | 432.9<br>(277.2) | 444.5<br>(303.7) | 457.3<br>(311.5) | 413.1<br>(278.7) | 416.3<br>(289.3) | 409.9<br>(288.1) |
| Labeled share |  | 52.0%<br>(18.1%) | 53.2%<br>(18.0%) | 52.9%<br>(18.0%) | 48.1%<br>(18.3%) | 50.0%<br>(18.7%) | 45.7%<br>(18.6%) |
| Saturated fat |  |  |  |  |  |  |  |
| Labeled |  | 35.1<br>(28.2) | 37.1<br>(31.4) | 39.6<br>(33.9) | 35.1<br>(31.2) | 37.1<br>(32.9) | 34.5<br>(32.0) |
| Non-labeled |  | 10.6<br>(12.9) | 10.7<br>(13.5) | 11.3<br>(14.4) | 11.8<br>(14.3) | 11.2<br>(13.6) | 13.2<br>(15.7) |
| Total |  | 45.7<br>(35.2) | 47.8<br>(38.9) | 50.9<br>(41.8) | 47.0<br>(38.9) | 48.2<br>(40.3) | 47.7<br>(41.3) |
| Labeled share |  | 77.3%<br>(18.8%) | 78.1%<br>(18.5%) | 78.2%<br>(18.4%) | 74.0%<br>(19.9%) | 76.0%<br>(19.2%) | 71.7%<br>(20.4%) |
| Sodium |  |  |  |  |  |  |  |
| Labeled |  | 412.7<br>(353.3) | 422.7<br>(379.6) | 429.1<br>(378.8) | 357.6<br>(324.9) | 349.1<br>(319.3) | 314.1<br>(306.9) |
| Non-labeled |  | 160.5<br>(190.9) | 159.0<br>(204.4) | 161.5<br>(204.2) | 157.7<br>(177.5) | 146.7<br>(176.7) | 159.4<br>(179.4) |
| Total |  | 573.2<br>(464.2) | 581.7<br>(500.6) | 590.6<br>(496.9) | 515.3<br>(428.8) | 495.8<br>(421.1) | 473.5<br>(412.9) |
| Labeled share |  | 72.4%<br>(19.2%) | 73.3%<br>(18.9%) | 72.9%<br>(19.1%) | 68.7%<br>(19.6%) | 70.0%<br>(19.5%) | 65.2%<br>(20.3%) |
| Sugars |  |  |  |  |  |  |  |
| Labeled |  | 85.5<br>(71.6) | 89.5<br>(77.2) | 87.5<br>(74.2) | 64.9<br>(59.5) | 66.1<br>(59.0) | 54.5<br>(51.8) |
| Non-labeled |  | 24.0<br>(22.5) | 23.4<br>(22.6) | 24.1<br>(23.1) | 28.2<br>(24.6) | 26.4<br>(23.3) | 32.1<br>(27.6) |
| Total |  | 109.4<br>(81.7) | 112.9<br>(86.9) | 111.6<br>(85.0) | 93.1<br>(71.5) | 92.5<br>(70.4) | 86.6<br>(66.3) |
| Labeled share |  | 75.9%<br>(16.9%) | 77.1%<br>(16.6%) | 76.0%<br>(17.0%) | 66.6%<br>(19.4%) | 68.4%<br>(19.0%) | 59.4%<br>(21.2%) |
| **Foods** |  |  |  |  |  |  |  |
|  |  | **2013-2014** | **2014-2015** | **2015-2016** | **2016-2017** | **2017-2018** | **2018-2019** |
| Energy |  |  |  |  |  |  |  |
| Labeled |  | 161.1<br>(118.8) | 167.3<br>(127.5) | 175.3<br>(134.2) | 154.1<br>(119.5) | 160.4<br>(126.1) | 149.1<br>(120.1) |
| Non-labeled |  | 171.5<br>(144.2) | 173.7<br>(158.5) | 180.3<br>(161.4) | 174.7<br>(154.3) | 175.0<br>(162.2) | 184.5<br>(165.3) |

|  |  |  |  |  |  |  |
| --- | --- | --- | --- | --- | --- | --- |
| Total | 332.7<br>(226.2) | 341.1<br>(249.2) | 355.6<br>(257.5) | 328.8<br>(235.3) | 335.4<br>(249.2) | 333.6<br>(249.6) |
| Labeled share | 50.7%<br>(20.7%) | 51.6%<br>(20.8%) | 51.7%<br>(20.7%) | 49.3%<br>(21.0%) | 51.0%<br>(21.5%) | 47.7%<br>(21.2%) |
| Saturated fat |  |  |  |  |  |  |
| Labeled | 34.2<br>(27.8) | 36.1<br>(30.9) | 38.5<br>(33.4) | 35.0<br>(31.1) | 36.2<br>(32.4) | 34.4<br>(31.9) |
| Non-labeled | 3.2<br>(5.9) | 3.3<br>(6.1) | 3.6<br>(6.5) | 4.3<br>(6.8) | 4.1<br>(7.2) | 5.6<br>(9.3) |
| Total | 37.4<br>(30.2) | 39.4<br>(33.4) | 42.2<br>(36.0) | 39.3<br>(33.9) | 40.3<br>(35.2) | 40.0<br>(36.3) |
| Labeled share | 90.7%<br>(12.4%) | 90.7%<br>(12.8%) | 90.5%<br>(12.8%) | 87.2%<br>(14.9%) | 88.9%<br>(14.3%) | 84.9%<br>(16.5%) |
| Sodium |  |  |  |  |  |  |
| Labeled | 393.2<br>(346.6) | 402.3<br>(372.5) | 409.4<br>(371.8) | 347.1<br>(321.7) | 337.1<br>(315.6) | 307.3<br>(305.5) |
| Non-labeled | 117.7<br>(173.8) | 116.8<br>(187.8) | 118.4<br>(186.3) | 113.0<br>(160.1) | 105.8<br>(160.9) | 115.0<br>(161.5) |
| Total | 510.9<br>(436.8) | 519.1<br>(471.9) | 527.8<br>(466.8) | 460.1<br>(404.6) | 442.9<br>(398.6) | 422.3<br>(391.5) |
| Labeled share | 78.5%<br>(20.6%) | 79.3%<br>(20.4%) | 78.8%<br>(20.5%) | 76.0%<br>(20.8%) | 77.2%<br>(20.6%) | 73.0%<br>(21.6%) |
| Sugars |  |  |  |  |  |  |
| Labeled | 32.8<br>(27.5) | 33.4<br>(28.2) | 33.8<br>(29.4) | 28.8<br>(26.3) | 31.1<br>(28.9) | 26.5<br>(25.5) |
| Non-labeled | 4.6<br>(5.2) | 4.6<br>(5.4) | 5.6<br>(6.2) | 7.1<br>(7.1) | 7.5<br>(7.6) | 10.5<br>(11.1) |
| Total | 37.4<br>(29.9) | 38.0<br>(30.7) | 39.4<br>(32.3) | 35.9<br>(29.6) | 38.6<br>(32.6) | 37.0<br>(31.5) |
| Labeled share | 84.8%<br>(15.0%) | 85.0%<br>(15.3%) | 82.3%<br>(16.8%) | 76.1%<br>(19.4%) | 76.4%<br>(19.4%) | 67.8%<br>(21.7%) |

Beverages
|  | 2013-2014 | 2014-2015 | 2015-2016 | 2016-2017 | 2017-2018 | 2018-2019 |
| --- | --- | --- | --- | --- | --- | --- |
| Energy |  |  |  |  |  |  |
| Labeled | 57.3<br>(62.3) | 61.0<br>(69.1) | 58.5<br>(64.7) | 37.6<br>(50.4) | 38.2<br>(51.6) | 28.8<br>(43.5) |
| Non-labeled | 42.9<br>(46.3) | 42.5<br>(47.8) | 43.2<br>(50.0) | 46.8<br>(50.4) | 42.8<br>(46.6) | 47.5<br>(50.0) |
| Total | 100.2<br>(84.3) | 103.4<br>(90.9) | 101.7<br>(89.6) | 84.3<br>(76.4) | 80.9<br>(74.1) | 76.3<br>(69.9) |
| Labeled share | 55.4%<br>(28.2%) | 57.3%<br>(28.1%) | 56.4%<br>(28.1%) | 42.1%<br>(29.4%) | 43.4%<br>(30.4%) | 34.0%<br>(30.6%) |
| Saturated fat |  |  |  |  |  |  |
| Labeled | 1.0<br>(1.9) | 1.0<br>(2.1) | 1.1<br>(2.2) | 0.2<br>(0.7) | 0.8<br>(5.2) | 0.1<br>(1.1) |
| Non-labeled | 7.4<br>(10.5) | 7.3<br>(11.0) | 7.6<br>(11.8) | 7.5<br>(11.4) | 7.1<br>(10.3) | 7.6<br>(11.1) |
| Total | 8.3 | 8.4 | 8.7 | 7.7 | 7.9 | 7.7 |
|  | (10.9) | (11.6) | (12.3) | (11.5) | (11.8) | (11.2) |
| Labeled share | 24.4% | 25.6% | 24.8% | 4.9% | 5.5% | 2.1% |
|  | (34.4%) | (35.2%) | (34.5%) | (16.9%) | (19.4%) | (11.3%) |
| Sodium |  |  |  |  |  |  |
| Labeled | 19.5 | 20.4 | 19.8 | 10.5 | 12.0 | 6.8 |
|  | (21.7) | (23.5) | (22.4) | (13.9) | (19.5) | (11.2) |
| Non-labeled | 42.8 | 42.2 | 43.1 | 44.7 | 40.9 | 44.4 |
|  | (46.9) | (47.3) | (49.4) | (48.0) | (44.3) | (47.0) |
| Total | 62.3 | 62.6 | 62.8 | 55.1 | 52.9 | 51.2 |
|  | (55.7) | (56.9) | (58.6) | (51.9) | (50.5) | (49.2) |
| Labeled share | 37.4% | 38.9% | 37.9% | 24.4% | 26.3% | 17.7% |
|  | (28.0%) | (28.2%) | (27.7%) | (24.4%) | (25.8%) | (22.9%) |
| Sugars |  |  |  |  |  |  |
| Labeled | 52.7 | 56.1 | 53.7 | 36.1 | 35.1 | 27.9 |
|  | (59.1) | (65.5) | (61.0) | (49.2) | (47.5) | (42.7) |
| Non-labeled | 19.3 | 18.9 | 18.5 | 21.1 | 18.9 | 21.6 |
|  | (20.3) | (20.3) | (20.3) | (21.4) | (19.5) | (21.6) |
| Total | 72.0 | 74.9 | 72.2 | 57.2 | 53.9 | 49.6 |
|  | (65.7) | (71.4) | (67.6) | (56.4) | (53.6) | (50.0) |
| Labeled share | 66.4% | 68.2% | 67.8% | 53.4% | 54.4% | 43.6% |
|  | (27.4%) | (27.0%) | (27.1%) | (31.2%) | (32.0%) | (33.8%) |

Energy, sugar, and saturated fat means are specified in terms of calories/capita/day purchased. Sodium is specified as mg/capita/day purchased.

**Supplemental Table 5.** Unadjusted weighted mean (SD) nutrient content of food and beverage purchases by policy period and SES.

| Total |  |  |  |  |  |  |
| --- | --- | --- | --- | --- | --- | --- |
|  | Pre-policy |  | Phase 1 |  | Phase 2 |  |
|  | High<br>SES | Low SES | High<br>SES | Low SES | High<br>SES | Low SES |
| Energy |  |  |  |  |  |  |
| Labeled | 263.7<br>(179.5) | 215.2<br>(151.4) | 216.5<br>(155.3) | 188.3<br>(136.6) | 190.5<br>(143.7) | 173.9<br>(128.8) |
| Non-labeled | 231.4<br>(186.7) | 213.8<br>(175.5) | 228.4<br>(180.5) | 216.8<br>(182.2) | 241.0<br>(194.6) | 229.2<br>(189.9) |
| Total | 495.2<br>(326.7) | 429.0<br>(286.6) | 445.0<br>(294.0) | 405.1<br>(280.1) | 431.4<br>(301.8) | 403.1<br>(283.3) |
| Labeled share | 54.7%<br>(16.5%) | 52.1%<br>(18.5%) | 50.0%<br>(17.5%) | 48.8%<br>(18.8%) | 45.8%<br>(18.0%) | 45.6%<br>(18.8%) |
| Saturated fat |  |  |  |  |  |  |
| Labeled | 46.4<br>(37.4) | 34.4<br>(28.5) | 44.4<br>(38.7) | 33.5<br>(29.2) | 42.8<br>(40.1) | 31.9<br>(28.5) |
| Non-labeled | 13.4<br>(15.7) | 10.0<br>(12.8) | 13.8<br>(16.0) | 10.8<br>(13.2) | 16.2<br>(19.6) | 12.2<br>(14.2) |
| Total | 59.8<br>(45.9) | 44.4<br>(35.5) | 58.2<br>(46.9) | 44.2<br>(36.3) | 59.0<br>(51.2) | 44.2<br>(36.9) |
| Labeled share | 77.6%<br>(17.9%) | 78.0%<br>(18.8%) | 75.3%<br>(18.8%) | 74.9%<br>(19.8%) | 71.1%<br>(20.3%) | 71.9%<br>(20.4%) |
| Sodium |  |  |  |  |  |  |
| Labeled | 464.3<br>(407.6) | 408.0<br>(357.4) | 375.2<br>(336.4) | 346.4<br>(317.1) | 310.3<br>(301.3) | 315.3<br>(308.6) |
| Non-labeled | 195.5<br>(227.0) | 149.1<br>(189.2) | 178.9<br>(197.1) | 143.6<br>(169.5) | 192.3<br>(210.0) | 149.1<br>(167.3) |
| Total | 659.8<br>(547.5) | 557.0<br>(464.2) | 554.2<br>(455.6) | 490.0<br>(413.7) | 502.6<br>(439.1) | 464.3<br>(403.9) |
| Labeled share | 71.1%<br>(18.3%) | 73.4%<br>(19.3%) | 67.3%<br>(19.0%) | 70.0%<br>(19.7%) | 60.9%<br>(20.2%) | 66.6%<br>(20.2%) |
| Sugars |  |  |  |  |  |  |
| Labeled | 91.9<br>(75.4) | 86.1<br>(74.0) | 63.7<br>(59.6) | 66.1<br>(59.1) | 49.5<br>(48.3) | 56.0<br>(52.8) |
| Non-labeled | 28.4<br>(26.3) | 22.4<br>(21.3) | 31.4<br>(27.1) | 25.9<br>(22.7) | 36.4<br>(32.0) | 30.8<br>(25.9) |
| Total | 120.2<br>(89.0) | 108.5<br>(82.9) | 95.2<br>(73.4) | 92.0<br>(70.1) | 85.9<br>(66.9) | 86.8<br>(66.1) |
| Labeled share | 74.6%<br>(16.9%) | 76.9%<br>(16.8%) | 63.9%<br>(19.7%) | 68.6%<br>(18.9%) | 54.8%<br>(21.5%) | 60.8%<br>(20.9%) |

Foods
|  | Pre-policy |  | Phase 1 |  | Phase 2 |  |
| --- | --- | --- | --- | --- | --- | --- |
|  | High SES | Low SES | High SES | Low SES | High SES | Low SES |
| Energy |  |  |  |  |  |  |
| Labeled | 206.2<br>(146.1) | 155.8<br>(118.0) | 184.8<br>(136.5) | 148.5<br>(116.8) | 169.3<br>(133.6) | 142.7<br>(114.7) |
| Non-labeled | 179.9<br>(159.3) | 173.7<br>(153.6) | 175.9<br>(153.2) | 174.5<br>(159.9) | 186.9<br>(163.3) | 183.8<br>(165.9) |
| Total | 386.1<br>(267.5) | 329.5<br>(235.7) | 360.6<br>(248.3) | 322.9<br>(239.7) | 356.3<br>(260.6) | 326.5<br>(245.5) |
| Labeled share | 55.4%<br>(18.8%) | 50.1%<br>(21.2%) | 53.2%<br>(19.7%) | 49.1%<br>(21.6%) | 49.6%<br>(20.2%) | 47.1%<br>(21.5%) |
| Saturated fat |  |  |  |  |  |  |
| Labeled | 45.2<br>(37.0) | 33.5<br>(28.1) | 44.0<br>(38.5) | 32.9<br>(28.8) | 42.7<br>(40.0) | 31.8<br>(28.4) |
| Non-labeled | 5.1<br>(8.3) | 2.9<br>(5.2) | 6.0<br>(9.6) | 3.6<br>(5.9) | 8.6<br>(13.8) | 4.7<br>(7.0) |
| Total | 50.3<br>(40.2) | 36.3<br>(30.1) | 50.0<br>(42.1) | 36.6<br>(31.1) | 51.3<br>(46.3) | 36.5<br>(31.7) |
| Labeled share | 89.1%<br>(13.1%) | 91.1%<br>(12.5%) | 86.6%<br>(15.0%) | 88.5%<br>(14.5%) | 81.7%<br>(17.9%) | 85.9%<br>(15.9%) |
| Sodium |  |  |  |  |  |  |
| Labeled | 442.9<br>(399.1) | 388.6<br>(351.0) | 365.0<br>(332.3) | 334.8<br>(313.9) | 305.2<br>(300.1) | 307.9<br>(307.1) |
| Non-labeled | 137.8<br>(204.4) | 111.2<br>(174.9) | 123.5<br>(175.8) | 104.9<br>(155.1) | 135.9<br>(185.1) | 108.5<br>(152.8) |
| Total | 580.7<br>(512.6) | 499.8<br>(438.6) | 488.4<br>(426.5) | 439.7<br>(392.8) | 441.1<br>(410.8) | 416.4<br>(385.1) |
| Labeled share | 78.2%<br>(19.7%) | 79.0%<br>(20.7%) | 75.9%<br>(20.2%) | 76.9%<br>(20.9%) | 69.7%<br>(21.5%) | 74.1%<br>(21.4%) |
| Sugars |  |  |  |  |  |  |
| Labeled | 40.2<br>(33.2) | 31.1<br>(26.3) | 34.2<br>(30.0) | 28.6<br>(26.7) | 29.2<br>(27.3) | 25.7<br>(24.9) |
| Non-labeled | 6.0<br>(6.8) | 4.6<br>(5.2) | 7.9<br>(7.9) | 7.1<br>(7.2) | 11.7<br>(13.1) | 10.1<br>(10.4) |
| Total | 46.2<br>(36.3) | 35.8<br>(28.6) | 42.1<br>(33.4) | 35.7<br>(30.3) | 40.8<br>(33.8) | 35.8<br>(30.6) |
| Labeled share | 84.5%<br>(14.6%) | 83.9%<br>(16.2%) | 77.4%<br>(18.1%) | 75.9%<br>(19.8%) | 68.3%<br>(21.3%) | 67.7%<br>(21.8%) |
| Beverages |  |  |  |  |  |  |
|  | Pre-policy |  | Phase 1 |  | Phase 2 |  |
|  | High SES | Low SES | High SES | Low SES | High SES | Low SES |
| Energy |  |  |  |  |  |  |
| Labeled | 57.5<br>(62.2) | 59.4<br>(66.4) | 31.8<br>(48.4) | 39.8<br>(51.7) | 21.1<br>(37.6) | 31.2<br>(45.0) |
| Non-labeled | 51.6<br>(54.3) | 40.1<br>(45.5) | 52.5<br>(53.8) | 42.3<br>(46.5) | 54.0<br>(56.4) | 45.4<br>(47.7) |
| Total | 109.1<br>(92.3) | 99.5<br>(86.9) | 84.3<br>(77.7) | 82.1<br>(74.5) | 75.1<br>(71.8) | 76.6<br>(69.3) |
| Labeled share | 51.8%<br>(28.4%) | 57.9%<br>(27.9%) | 34.4%<br>(29.5%) | 45.4%<br>(29.6%) | 24.6%<br>(28.6%) | 36.9%<br>(30.6%) |
| Saturated fat |  |  |  |  |  |  |
| Labeled | 1.3<br>(2.6) | 0.9<br>(1.9) | 0.4<br>(3.1) | 0.5<br>(3.9) | 0.1<br>(1.0) | 0.1<br>(1.1) |
| Non-labeled | 8.3<br>(12.0) | 7.2<br>(10.8) | 7.8<br>(11.4) | 7.1<br>(10.7) | 7.6<br>(11.7) | 7.5<br>(10.9) |
| Total | 9.6<br>(12.5) | 8.1<br>(11.3) | 8.2<br>(11.9) | 7.7<br>(11.5) | 7.7<br>(11.7) | 7.7<br>(11.0) |
| Labeled share | 24.3%<br>(34.1%) | 25.1%<br>(34.9%) | 4.9%<br>(17.7%) | 5.3%<br>(18.4%) | 1.7%<br>(10.0%) | 2.2%<br>(11.7%) |
| Sodium |  |  |  |  |  |  |
| Labeled | 21.4<br>(24.8) | 19.4<br>(21.7) | 10.3<br>(17.8) | 11.5<br>(16.7) | 5.1<br>(10.3) | 7.3<br>(11.4) |
| Non-labeled | 57.8<br>(59.8) | 37.9<br>(42.3) | 55.4<br>(55.5) | 38.8<br>(42.0) | 56.4<br>(58.7) | 40.6<br>(42.0) |
| Total | 79.1 | 57.3 | 65.7 | 50.3 | 61.6 | 47.9 |
|  | (69.5) | (51.4) | (60.3) | (47.3) | (60.2) | (44.7) |
| Labeled share | 32.6% | 39.8% | 19.0% | 27.4% | 11.5% | 19.7% |
|  | (26.4%) | (28.2%) | (22.4%) | (25.6%) | (19.0%) | (23.7%) |
| Sugars |  |  |  |  |  |  |
| Labeled | 51.7 | 55.0 | 29.6 | 37.5 | 20.4 | 30.3 |
|  | (57.7) | (63.3) | (45.6) | (49.0) | (36.3) | (44.2) |
| Non-labeled | 22.3 | 17.8 | 23.5 | 18.9 | 24.8 | 20.7 |
|  | (23.4) | (19.1) | (23.6) | (19.3) | (25.1) | (20.3) |
| Total | 74.0 | 72.7 | 53.1 | 56.3 | 45.1 | 51.0 |
|  | (67.0) | (68.7) | (54.3) | (55.2) | (46.7) | (50.9) |
| Labeled share | 63.5% | 68.8% | 44.7% | 56.8% | 32.5% | 47.2% |
|  | (28.3%) | (26.7%) | (32.6%) | (30.7%) | (33.2%) | (33.3%) |

**Supplemental Table 6.** Weighted adjusted mean differences between the observed post-policy purchases and counterfactual purchases of calories from total foods and beverages across models using different pre-policy windows.

| #<br>months<br>pre-policy | Policy time-window | Phase 1 |  |  |  |  |  | Phase 2 |  |  |  |  |  |
| --- | --- | --- | --- | --- | --- | --- | --- | --- | --- | --- | --- | --- | --- |
|  |  | Total |  | Labeled |  | Non-labeled |  | Total |  | Labeled |  | Non-labeled |  |
|  |  | Absolute<br>(95%<br>CI) | Relative<br>(95%<br>CI) | Absolute<br>(95%<br>CI) | Relative<br>(95%<br>CI) | Absolute<br>(95%<br>CI) | Relative<br>(95%<br>CI) | Absolute<br>(95%<br>CI) | Relative<br>(95%<br>CI) | Absolute<br>(95%<br>CI) | Relative<br>(95%<br>CI) | Absolute<br>(95%<br>CI) | Relative<br>(95%<br>CI) |
| 42 | January 2013-June 2016 | **-36.9**<br>(-44.9,-29.0) | **-8.2**<br>(-9.8,-6.5) | **-39.3**<br>(-43.9,-34.6) | **-16.8**<br>(-18.5,-15.2) | **2.3**<br>(-2.4,7.0) | **1.1**<br>(-1.1,3.3) | **-30.7**<br>(-44.9,-16.5) | **-7.1**<br>(-10.2,-4.1) | **-50.3**<br>(-58.3,-42.3) | **-22.5**<br>(-25.4,-19.7) | **19.5**<br>(10.9,28.2) | **9.4**<br>(4.9,13.8) |
| 36 | July 2013- June 2016* | **-40.0**<br>(-48.5,-31.5) | **-8.8**<br>(-10.5,-7.1) | **-40.0**<br>(-45.1,-34.9) | **-17.1**<br>(-18.9,-15.3) | **-0.1**<br>(-5.2,5.0) | **0.0**<br>(-2.4,2.3) | **-36.2**<br>(-51.8,-20.6) | **-8.3**<br>(-11.6,-5.0) | **-51.6**<br>(-60.7,-42.6) | **-23.0**<br>(-26.2,-19.8) | **15.1**<br>(5.6,24.6) | **7.1**<br>(2.3,11.9) |
| 30 | January 2014- June 2016 | **-33.6**<br>(-42.8,-24.4) | **-7.5**<br>(-9.4,-5.5) | **-36.0**<br>(-41.5,-30.6) | **-15.6**<br>(-17.6,-13.6) | **2.4**<br>(-3.2,8.0) | **1.1**<br>(-1.5,3.7) | **-24.8**<br>(-41.6,-8.0) | **-5.9**<br>(-9.6,-2.1) | **-44.6**<br>(-54.3,-34.9) | **-20.5**<br>(-24.1,-16.9) | **19.7**<br>(9.4,30.0) | **9.4**<br>(4.1,14.8) |
| 24 | July 2014- June 2016 | **-33.1**<br>(-42.8,-23.5) | **-7.4**<br>(-9.4,-5.3) | **-34.4**<br>(-40.0,-28.8) | **-15.0**<br>(-17.1,-12.9) | **1.2**<br>(-4.7,7.1) | **0.6**<br>(-2.1,3.3) | **-23.8**<br>(-41.4,-6.2) | **-5.6**<br>(-9.5,-1.7) | **-41.9**<br>(-51.9,-31.8) | **-19.5**<br>(-23.3,-15.7) | **17.8**<br>(7.0,28.6) | **8.4**<br>(2.9,14.0) |
| 18 | January 2015-June 2016 | **-17.0**<br>(-27.7,-6.3) | **-3.8**<br>(-6.2,-1.4) | **-25.7**<br>(-31.9,-19.5) | **-11.5**<br>(-14.1,-9.0) | **8.8**<br>(2.2,15.3) | **4.2**<br>(0.9,7.5) | **6.6**<br>(-12.9,26.0) | **1.6**<br>(-3.4,6.6) | **-25.7**<br>(-36.6,-14.8) | **-12.9**<br>(-17.7,-8.1) | **32.2**<br>(20.1,44.3) | **16.4**<br>(9.3,23.5) |

\* This is the main model used in the paper

**Supplemental Table 7.**
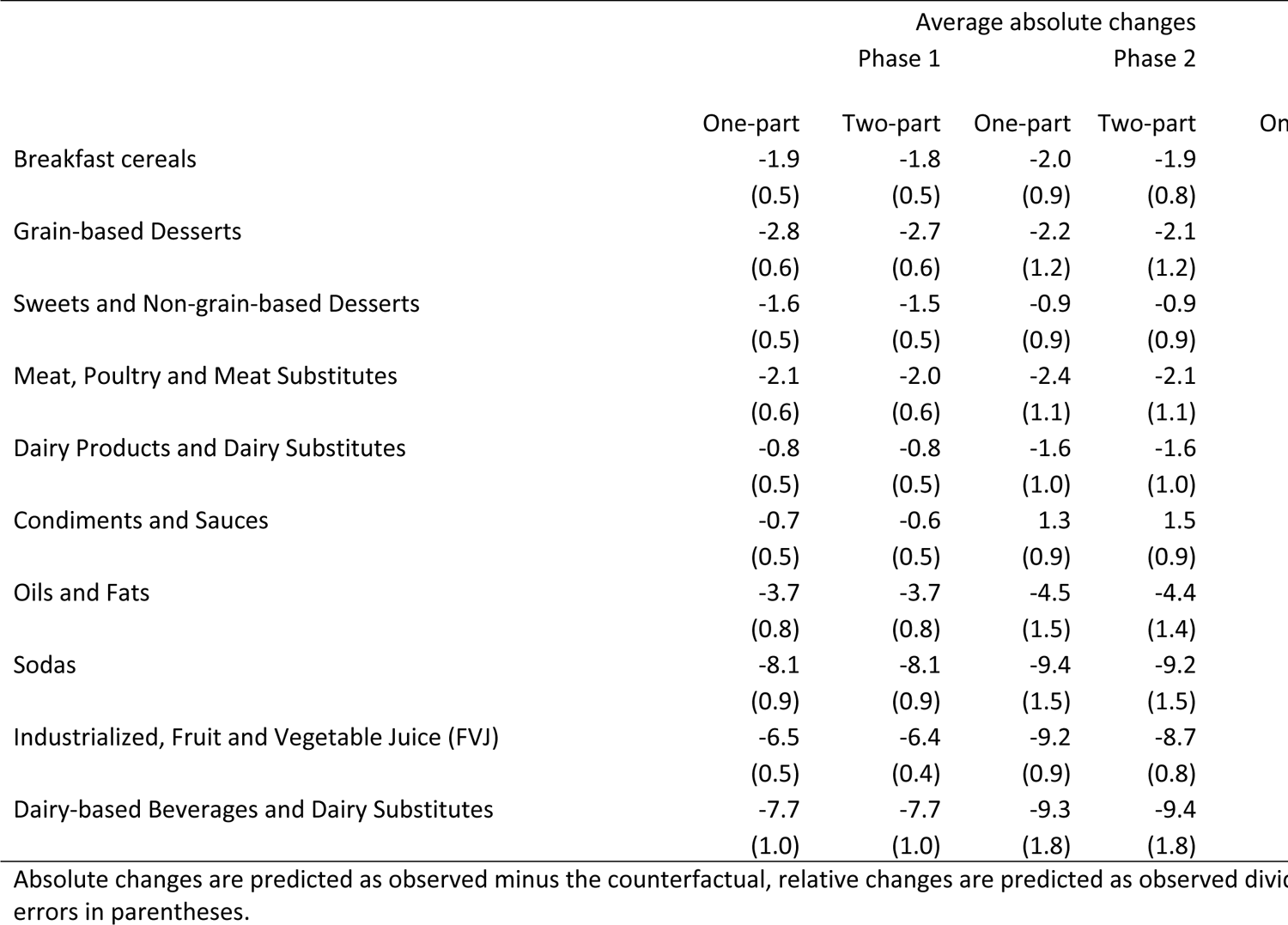
One-part vs. two-part models for total calories (Labeled and Non-labeled) by food and beverage group.

**Supplemental Table 8.** Differences between papers

The overall pattern of results reported in the current study is consistent with our previously published evaluations of the first phase of the Chilean Law using Kantar WorldPanel data in that they show an overall decline in calories driven by a decline in Labeled calories, with partial compensation by an increase in Non-labeled calories. However, when comparing current Phase 1 results for total food and beverage purchases to our previously published results, there are several differences. Compared to our previously published results, declines in Labeled calories purchased are smaller and increases in Non-labeled calories purchased are also smaller, resulting in a larger estimation of declines in overall calories purchases. This pattern is consistent across nutrients, with the exception that for sugar, current results yield a larger decline in Labeled calories from sugar than did previous rsesults. There are multiple explanations for these findings relating to differences in the datasets used for these analyses. In our previous results, we applied Phase 1 cutoffs to classify Labeled vs. Non-labeled foods, whereas in our current results, we applied Phase 3 (more stringent) cut-offs across all phases in order to have a consistent definition of regulation status over time; these differences in classifications resulted in a higher proportion of foods being classified as “Labeled” and less as “not high in” during Phase 1 in our current analysis. In our previous results, we used a shorter pre-policy window (January 1, 2015-June 2016) whereas in our current results, we used a longer pre-policy window (July 1, 2013-June 2016) in order to achieve balance with the longer post-policy window, since we were examining both Phase 1 and Phase 2. As shown in the sensitivity results, altering the pre-policy window changes the counterfactual, which is based on pre-existing trends, and can lead to estimation of larger or smaller differences between the observed and counterfactual. Similarly, our current results included the entire Phase 1 (July 1, 2016-June 30, 2018), and included updated Nutrition Facts Panel information collected in the first semester of 2018, whereas our previous results were truncated at December 31, 2017 due to data availability at the time of publication. It is possible that products were additionally reformulated in 2018 in anticipation of Phase 2 of the law, which was implemented in July 2018 and included stricter nutrient thresholds, which would likely lead to larger reductions in nutrients purchased. Finally, there were other differences between the analyses, including covariates (e.g., public holidays by region and month, inclusion of the SES variable), weighting technique (matching by production period vs. month), and model specifications, which reflect our updated understanding of the dataset. The differences in results highlights the need for careful attention to methodological issues such as how products are reviewed and linked to NFP data as well as how pre- and post-policy time windows are created. As policy implementation continues across the globe, it will be important to understand not only how policy-linked purchasing patterns change over time, but how to develop and implement methodological strategies for natural experiments to evaluate these.

